# Gene Cluster Expression Index (GCEI) and Potential Indications for Targeted Therapy and Immunotherapy

**DOI:** 10.1101/2023.06.17.23291532

**Authors:** Aibing Rao

## Abstract

Lung cancer recurrence risk was demonstrated to be related to driver gene and immunotherapy target gene cluster expression abnormality. Nine clusters seeded with driver genes ALK, BRAF, EGFR, MET, NTRK, RAS, RET, ROS1, TP53 and two immunotherapy target genes PDCD1 and CTLA4 were investigated respectively to predict lung cancer recurrence. The cluster of a seed was pre-selected to include fusion partner genes in the case of gene fusion, ligands, its pseudogenes, upstream and downstream co-expressors or inhibiting genes, effectors directly related to important pathways, etc. For each cluster, a gene cluster expression index (GCEI) was defined in two steps: Firstly, apply the univariate ROC of using each member’s expression vector to predict recurrences to label a patient sample as either normal or abnormal; Secondly, apply the percentage of abnormal genes in the cluster to predict recurrences to derive an optimal threshold so that a cluster member voting strategy can be achieved and a sample is labeled as abnormal (with respect to the cluster expression profile) if the the percentage of abnormal genes for the sample is greater than or equal to the threshold and as normal vice versa. Combinatory GCEI was developed as a binary string concatenating the individual GCEI corresponding to the individual cluster in an ordered list of driver or other important gene seeds. It showed that the recurrence risk of the abnormal group is typically 50% to 200% higher than the normal counterpart. Finally it was proposed and discussed to expand targeted therapy and immunotherapy to the abnormal group defined by GCEI.

**Background:** Molecular profiling such as DNA-based mutation panels and proteiomics have been demonstrated great success in oncology for personalized medicine. Transcriptome profiling has emerged to be another promising opportunity as complement and expansions to the DNA-based approach and as new tools to further advance clinical oncology.

**Methods:** Lung cancer gene expression GEO data sets were downloaded, normalized, combined and analyzed. A novel approach was presented to analyze expression abnormality of important gene clusters with seeds including drivers such as ALK, BRAF, EGFR, MET, NTRK, RAS, RET, ROS1, TP53 or immunotherapy target PDCD1 and CTLA4, etc. A cluster was pre-specified for each seed and included the fusion partners in the case of translocation, ligands, activators, inhibitors, effectors, co-stimulators in the important pathways, etc. Each cluster member was labeled as normal or abnormal (up or down) with the univariate ROC by using its expression to predict recurrences. Cluster level labeling of expression state (normal or abnormal) was via a dynamic voting strategy, of which the voting threshold was set as the optimal cutoff on the ROC associated with the univariate model of using the percentage of the abnormal members to predict recurrences. Given an ordered list of important genes, a binary string of the same length was encoded by assigning 0 for *normal* and 1 for *abnormal* representing the cluster expression state of the corresponding position, called gene cluster expression index (GCEI) signature. Finally lung cancer recurrences were assessed and compared based on GCEI states and the combinations.

**Results:** The recurrence risks of single gene normal group (*GCEI* = 0) vs abnormal group (*GCEI* = 1) were as follows, ALK: 17% vs. 55% for all stages, 13% vs. 42% for Stage I, 36% vs. 67% for Stage II-IV; BRAF: 23% vs. 49% for all stages, 15% vs. 36% for Stage I, 54% vs. 59% for Stage II-IV; EGFR: 25% vs. 47% for all stages, 17% vs. 33% for Stage I, 54% vs. 59% for Stage II-IV; MET: 25% vs. 44% for all stages, 17% vs. 29% for Stage I, 51% vs. 60% for Stage II-IV; NTRK: 19% vs. 52% for all stages, 13% vs. 40% for Stage I, 44% vs. 63% for Stage II-IV; RAS: 24% vs. 51% for all stages, 16% vs. 35% for Stage I, 47% vs. 65% for Stage II-IV; RET: 19% vs. 50% for all stages, 14% vs. 35% for Stage I, 40% vs. 65% for Stage II-IV; ROS1: 23% vs. 48% for all stages, 17% vs. 32% for Stage I, 45% vs. 64% for Stage II-IV; TP53: 23% vs. 50% for all stages, 15% vs. 38% for Stage I, 49% vs. 64% for Stage II-IV; and for the immunotherapy target gene: CTLA4: 26% vs. 49% for all stages, 14% vs. 38% for Stage I, 53% vs. 62% for Stage II-IV; PDCD1: 28% vs. 48% for all stages, 16% vs. 37% for Stage I, 54% vs. 61% for Stage II-IV. In addition, taking 9-driver gene GCEI and summarizing number of ‘1’, the count of abnormal driver genes, *N*, and then comparing the population of *N ≤* 5 vs. *N >* 5, the recurrence risks were: 19% vs. 59% for all stages, 13% vs. 49% for Stage I, 41% vs. 66% for Stage II-IV. Hence most of the cases the recurrence risk is 1.5 to 3 times higher for patient group with abnormally expressed gene clusters than normally expressed.

**Discussion:** Precision medicine based on RNA expression analysis is discussed and it is conjectured to apply targeted therapy or immunotherapy to lung cancers based on the related gene expression status as determined by the cluster member voting strategy. This can serve as an extension and complement to the current DNA-based tests, especially for a majority of patients who have been tested negative based on the conventional tests and have possibly missed the potential treatment benefit.

## 1 Introduction

Gene expression or transcriptome profiling has been extensively explored in the past 20 years in oncology and there are several multi-gene Rna tests have been put in practical clinical use for human cancers[44, 46, 58]. For lung cancer there have been a lot of gene expression signatures published for prognosis prediction[7, 9, 26, 29, 33, 63, 64, 67, 71] and a comprehensive evaluation was performed by Tang et al.[59] However there is little research regarding using gene expression profiling for targeted therapy or immunotherapy. The current standard approach for selection of targeted therapy is via matching particular gene mutations[39], and the selection of immunotherapy is via routine tests such as pathological immunoassay (IHC) for protein expression of PD1 or PD-L1, via DNA-based NGS assessment of Tumor Mutation Burden (TMB), Mismatch Repair (MMR), and Micro-satellite Instability (MSI). Transcriptome profiling analysis has emerged as promising biomarkers to cancer treatment and showed encouraging clinical results[10, 57]. In NSCLC, study showed that gene expression profiling might have better prognostic prediction power than mutation status[38] in particular scenarios. Hence, RNA profiling analysis is promising and will be an important direction as a complement or even better choice than the current IHC and DNA-based approaches in precision medicine of cancers. In the following we present a framework with novel transcriptome analysis algorithms to assign expression abnormality status to important driver genes or immunotherapy target genes based on member smart voting within a clustered gene set seeded at considering gene for lung cancer. The patient populations in different expression state have been showed to have dramatically different clinical prognosis risks and hence requires different personalized treatment considerations. The general framework is applicable and implementable to other cancer types with strongly related genes with clusters.

## 2 Materials and methods

### 2.1 Gene Expression Data

Two microarray datasets downloaded from Gene Expression Omnibus (GEO) databases are GSE30219 [48] and GSE31210 [40]. There were 483 lung cancers with none-empty recurrence labels, of which 204 cases were labeled as recurred within two years since the diagnosis, accounting for 42%. Two data sets were respectively normalized using IQR (Inter Quantile Range) method, namely, the the quartiles (*Q*1, *Q*3) of the original expression data were linearly mapped to the unit interval (0,> 1). The normalization procedure was applied first at the sample dimension and then at the gene dimension. Then taking the common genes, two normalized data subsets of the common genes plus clinical variables were stacked together to form a combined analysis data set. There are about 17000 common genes, the cases of different stages counted as 310 (I), 111 (II), 44 (III-IV), and 8 (unknown). Average age is 61, with the youngest 15 and the eldest 84. There are 331 male patients.

### 2.2 Pre-selected Gene Clusters

Nine driver gene ALK, BRAF, EGFR, MET, NTRK, RAS, RET, ROS1, TP53 and two immunotherapy target genes PDCD1, CTLA4 were used for gene cluster expression analysis. Take ALK as an example, in ALK-positive NSCLC population, almost 100 ALK fusion partners were cataloged [41], together with String and genecard.org description, 107 genes were selected. The gene clusters are listed in Table 1. Note that the clusters are not mutually exclusive and one member may appear in different clusters.

**Table 1:**
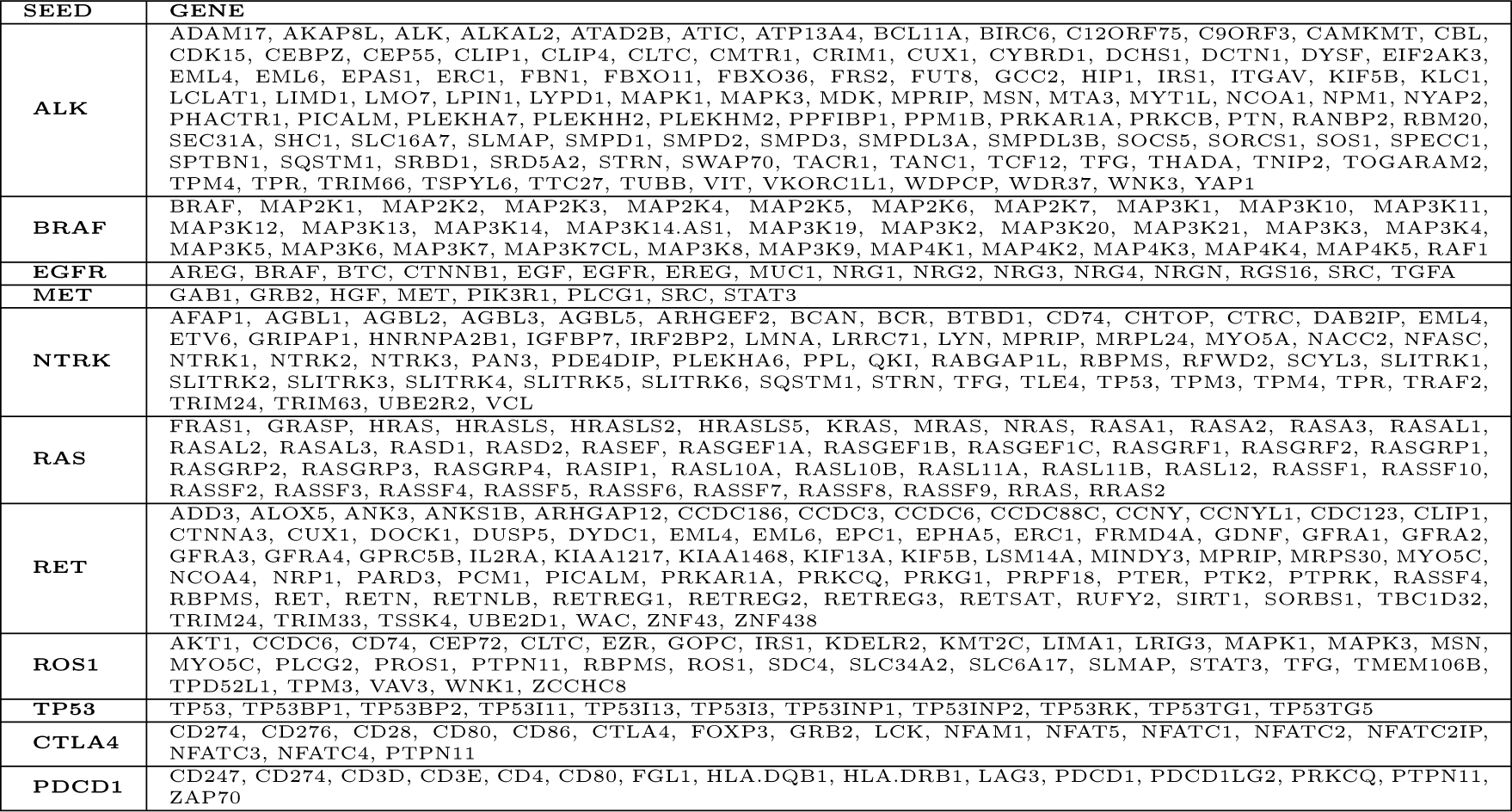
Pre-selected Gene Clusters for Important Lung Cancer Genes.

### 2.3 Gene Cluster Expression Index (GCEI)

The goal is to assign a sample a binary index for a given gene cluster, called *gene cluster expression index* (GCEI). It consists of two steps, first to determine the expression index of all cluster members, second to apply a smart voting procedure to define the GCEI for the sample.

#### 2.3.1 ROC of Univariate Model to Determine Single Gene Expression Abnormality with Respect to Recurrence

Given a gene seed, use each cluster member listed in Table 1 to predict recurrence and draw ROC to obtain an optimal cutoff, which is set at the ROC position closest to the top-left corner of the unit square, the cutoff is used to determine a sample expression status: normal or abnormal. Given a member gene *g*, let *T_g_* be the cutoff, then the samples in the training data set are divided into two populations, one above *T_g_*, the other below. Then for each population, the recurrence percentages are collected, denoted as *P_above_, P_below_*,respectively. Let *P_δ_*= *|P_above_ − P_below_|*, the absolute difference values between the populations, it represents the prediction power of a gene expression to recurrence. *P_δ_* represents the prediction power of *g* as a univariate predictor of the recurrence. Moreover, if *P_above_ > P_below_*, then *g* is over-expressed for the population of higher recurrence risk, or else if *P_above_ < P_below_*, then *g* is down-expressed. The risk difference between these two groups is called *significant* if *P_δ_ ≥ T_diff_*, where *T_diff_* is a pre-specified threshold and is set as 5% in the following. With respect to *g*, a sample is labeled as: (1). *noraml* if *P_δ_ < T_diff_*; (2). *up* if *P_δ_ ≥ T_diff_* and *P_above_ > P_below_*; (3). *down* if *P_δ_ ≥ T_diff_* and *P_above_ < P_below_*. Both *up* and *down* are called *abnormal*.

#### 2.3.2 Cluster Member Voting to Define GCEI

Now for the considering cluster, calculate the percentage of the *abnormal* gene members for each sample and use the percentage as a new univariate predictor of recurrence, following the same approach as in the above, ROC is plotted and an optimal percentage threshold *T_p_* is obtained. Now for each sample, if the percentage of the abnormal members is greater than or equal to *T_p_*, the sample is labeled with 1, or else 0. This characteristic index is called *Gene Cluster Expression Abnormality Index* (GCEI). GCEI value 1 represents *abnormal* expression with respect to the given gene cluster, while value 0 represents *normal*.

#### 2.3.3 Lung Cancer Recurrence Risks of GCEI Status

Recurrence risks are assessed with respect to the status of a single GCEI or a combination of multiple GCEIs. For a single cluster GCEI, recurrence risk is calculated for *GCEI* = 0 and *GCEI* = 1 respectively. For 9 driver gene cluster combination, given the ordered list (ALK, BRAF, EGFR, MET, NTRK, RAS, RET, ROS1, TP53), concatenate the corresponding GCEI of each cluster to obtain a binary string of 9 bits, for example, 000000000 represents all nine gene clusters are normally expressed, 100000000 represents only ALK cluster is abnormally expressed, 111111111 represents all 9 clusters are abnormally expressed, and so on. 9-bit GCEI classifies lung cancers into 2^9^ = 512 subtypes. Moreover, since in practice it might be difficult to accumulate enough patient cases for some of the 512 subtypes, we may collapse 512 subtypes into only 10 super-subtypes as follows, by counting number of digit 1 in the 9-bit string, patients are grouped into 10 subtypes with aggregated GCEI of 0,> 1,> 2,> 3, *· · ·,* 9 respectively, and each GCEI value tells how many gene clusters are abnormal among the nine clusters. For immunotherapy target couple (CTLA4, PDCD1), GCEI is a two-digit string with four combinations: 00,> 01,> 10,> 11, representing none, CTLA4 only, PDCD1 only, or both of the two clusters are abnormally expressed respectively.

### 2.4 Data Analysis Software

Data analysis and plots were scripted in house using RStudio 2022.07.1 with R version 4.0.5 on Mac platform with OS version darwin17.0.

## 3 Results

### 3.1 Univariate Models

Univariate models are constructed first by using the expression of each cluster member as a recurrence predictor and second by aggregating the expression status of cluster members and using the percentage of abnormal members as a new recurrence predictor, called cluster member voting model. At last, recurrence risks are assessed with respect to various patient populations using these models with combination.

#### 3.1.1 ALK Cluster

There are 107 pre-selected members in ALK cluster, most of which are fusion partners [41]. The univariate models showed that 72 *abnormal* genes have *P_δ_ ≥* 5%, accounting for 67%, and the rest 35 *normal* genes have *P_δ_ <* 5%. The corresponding AUCs, FPRs, TPRs, threshold *T_g_*, population risks of the *abnormal* and the *normal* genes are listed in Table 2 and Table 3 respectively. As shown in Table 2, 33 genes are over-expressed for higher recurrence risk: CEP55, TUBB, MDK, NPM1, CEBPZ, TFG, ATIC, LYPD1, LCLAT1, LPIN1, MYT1L, WNK3, TNIP2, C12ORF75, TPM4, TTC27, SOS1, ADAM17, TSPYL6, KLC1, PPFIBP1, SPECC1, FRS2, SHC1, FBN1, THADA, SQSTM1, CLIP1, CBL, CLTC, FBXO36, FUT8 and ITGAV; 39 are down-expressed for higher recurrence risk: ATP13A4, LMO7, WDR37, EPAS1, GCC2, CRIM1, PLEKHH2, TRIM66, FBXO11, SMPD1, YAP1, MPRIP, TANC1, SEC31A, PRKAR1A, CYBRD1, SPTBN1, ALKAL2, WDPCP, SLMAP, CLIP4, SLC16A7, SWAP70, LIMD1, BIRC6, SOCS5, PLEKHA7, EIF2AK3, PPM1B, KIF5B, PHACTR1, CAMKMT, RBM20, SRD5A2, NYAP2, PTN, PICALM, VKORC1L1 and HIP1.

**Table 2:**
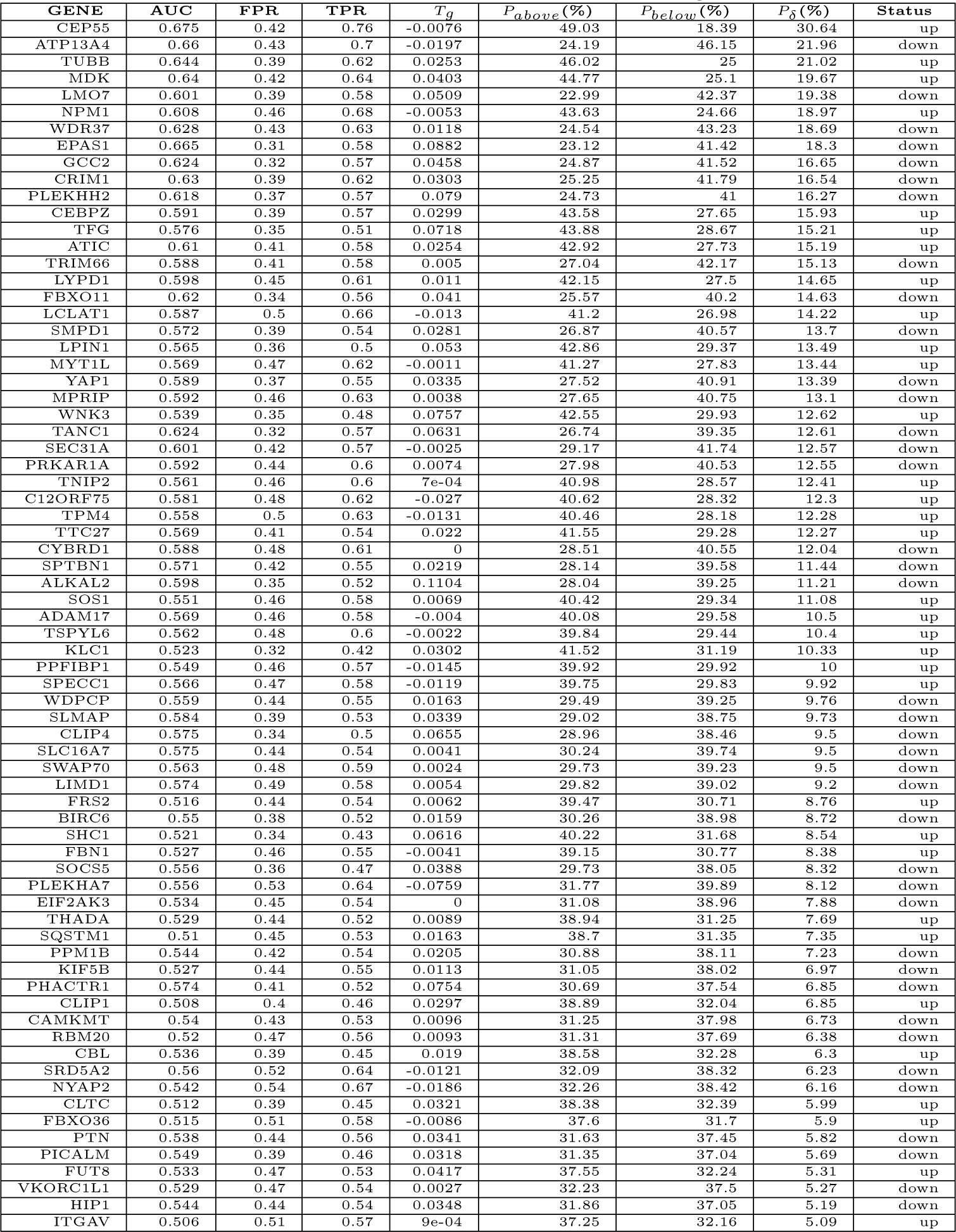
AUCs and recurrence risks of 72 *abnormal* ALK genes with *P_δ_ ≥* 5%.

**Table 3:**
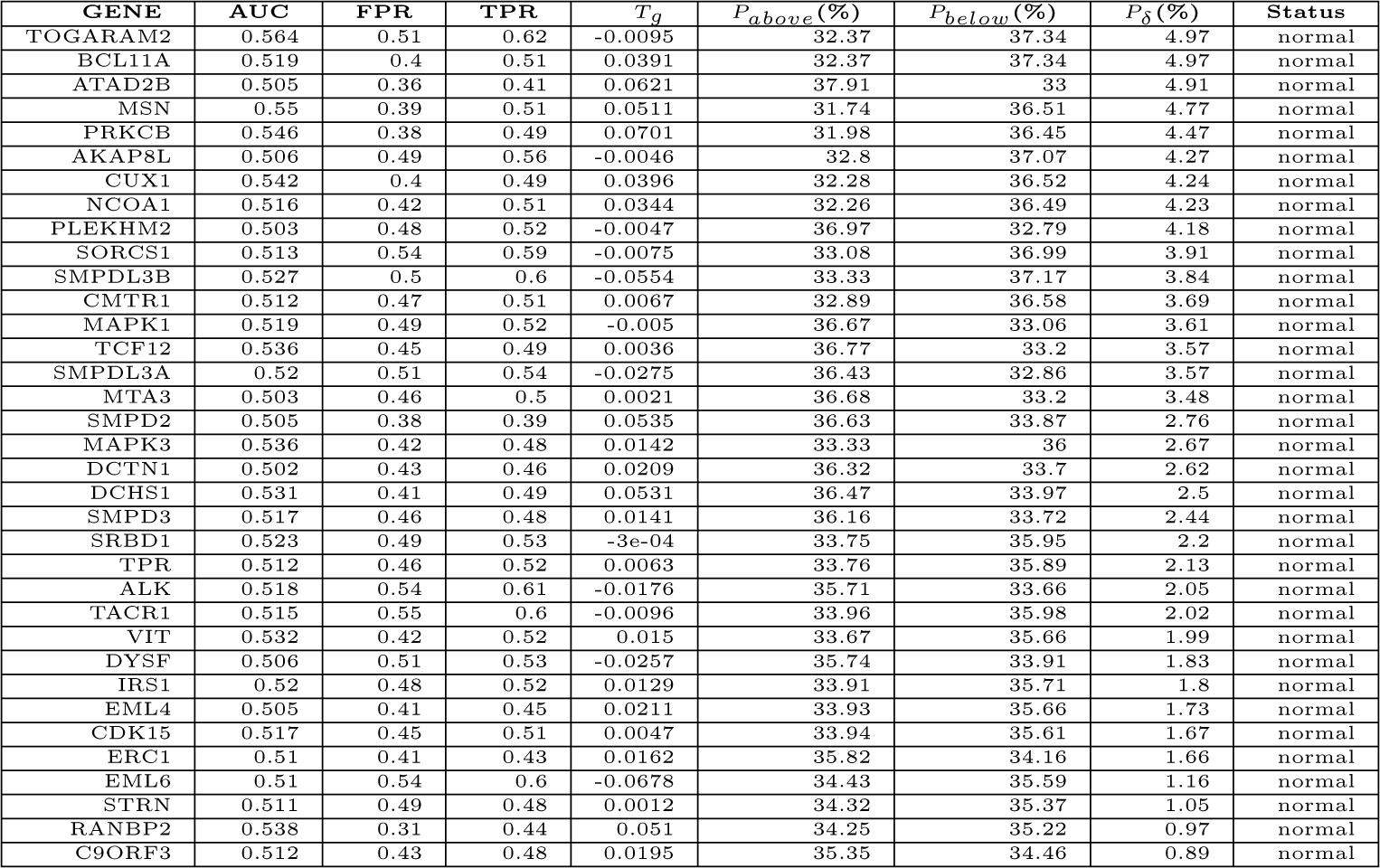
AUCs and recurrence risks of 35 *normal* ALK genes with *P_δ_ <* 5%.

For illustrating purpose, the ROCs of top 12 genes in decreasing order of *P_δ_* are shown in Figure 1. The highest one in the first row of Table 2 is CEP55. It shows that the normalized expression cutoff *T_g_* = *−*0.0076. Patience with CEP55 expression *≥* (*−*0.0076) has a recurrence risk of *P_above_* = 49.03% while patience with CEP55 expression *<* (*−*0.0076) has a risk of *P_below_* = 18.39%, hence the difference *P_δ_* = 30.64%. CEP55 is over-expressed (with respect to recurrence) because *P_above_ > P_below_*. CEP55, called Centrosomal Protein 55, is related to DNA damage and cytoskeletal signaling and plays a role in mitotic exit and cytokinesis. CEP55 was found to be a fusion partner of ALK [13] and high CEP55 expression is associated with poor prognosis [25]. The second gene is ATP13A4, which is down-expressed with *P_above_* = 46.15%, *P_below_*= 24.19% and a difference *P_δ_* = 21.96%. ATP13A4, called ATPase 13A4, may enable ATPase-coupled cation transmembrane transporter activity and may be involved in cellular calcium ion homeostasis.

**Figure 1:**
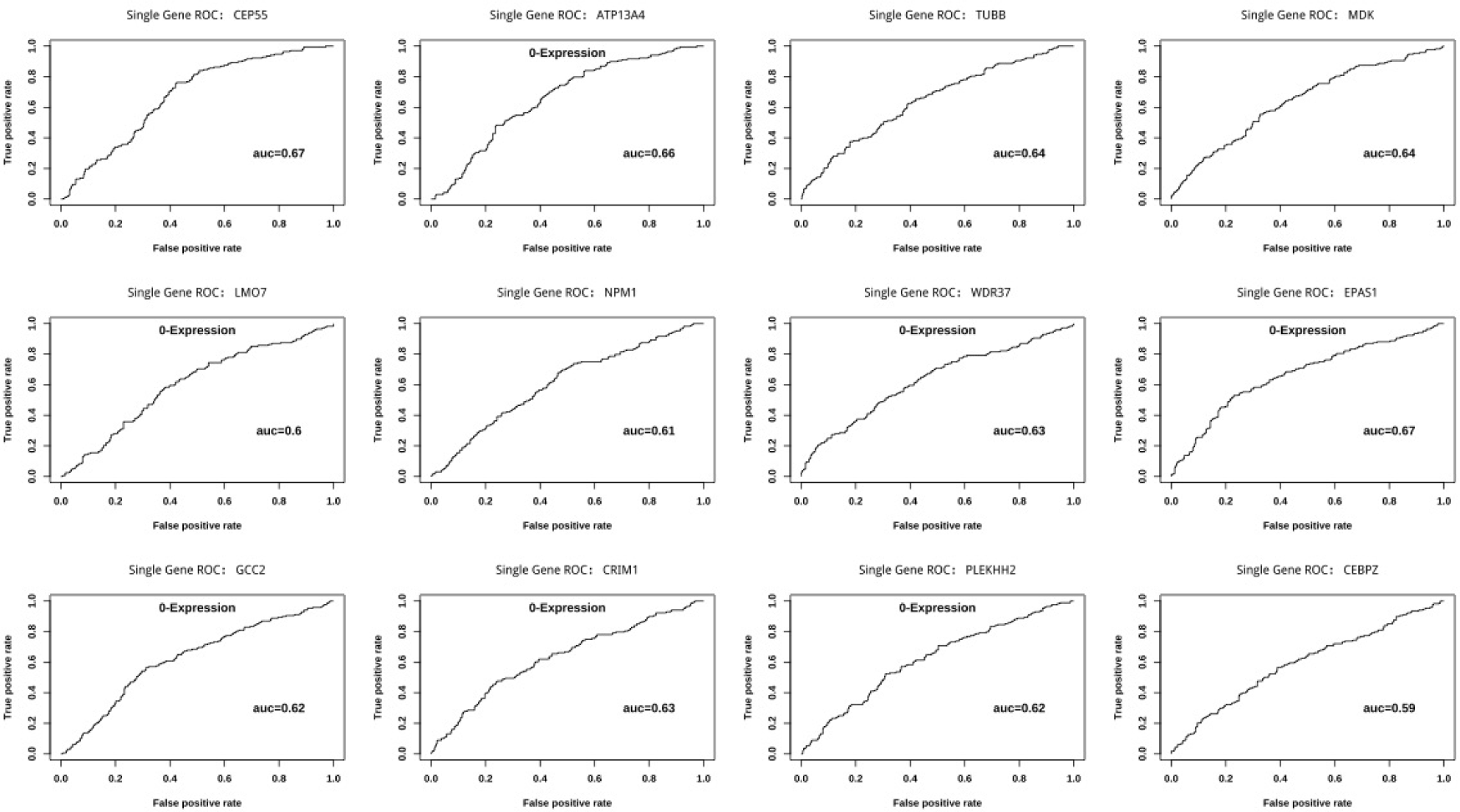
Univariate ROCs of the top 12 genes ALK cluster in the decreasing order of *P_δ_*. Inverted expression value (0 *− Expression*) was used to plot ROC for down regulated genes, similarly hereinafter.

In a lung cancer case study [11], a 53-year-old metastatic Stage IV patient was harbored with ATP13A4-ALK and two other ALK-fusions COX7A2L-ALK and LINC01210-ALK, firstline crizotinib therapy showed 12 months of PFS/PR, then a new SLCO2A1-ALK fusion led to resistance, afterwards a second line ceritinib resulted in further 8 months of PFS and NGS results demonstrated the loss of ATP13A4-ALK and SLCO2A1-ALK.

Interestingly, ALK expression itself is normal and only gives a difference of *P_δ_*= 2.02% with this training data set.

#### 3.1.2 BRAF Cluster

BRAF cluster contains 33 members. The ROCs of only 12 genes with top *P_δ_* are presented in Figure 2. AUCs, FPRs, TPRs, threshold *T_g_*, population risks for all members are listed in Table 4. BRAF phosphorylates MAP2K1 and thereby activates the MAP kinase signal pathway and here most of the cluster members are related to MAP. There are 19 members with *P_δ_ ≥* 5%, accounting for 58%, within which only 4 genes MAP2K2, MAP4K4, MAP3K7 and MAP2K1 are over-expressed. MAP2K1 (MEK1) and MAP2K2 (MEK2) activates BRAF via controlling KSR1[28]. On the other hand, RAF1 is down-expressed with modest *P_δ_* = 6.02%. BRAF/RAF1 heterodimers are downstream receptors of RAS and are crucial activator of MAPK[62]. Moreover, MAP2K3/4/5 and MAP3K1/2/3/5/6/7CL/8/9/11/13/14-AS1 are down-expressed with *P_δ_*ranging from 5.2% to 13.78%, so is RAF1. Other remaining ones such as MAP2K6/7, MAP3K/4/10/12/14/19/20 and MAP4K1/2/3/5 are normal with *P_δ_ <* 5%. BRAF itself is deemed to be normal with *P_δ_* = 4.21%.

**Figure 2:**
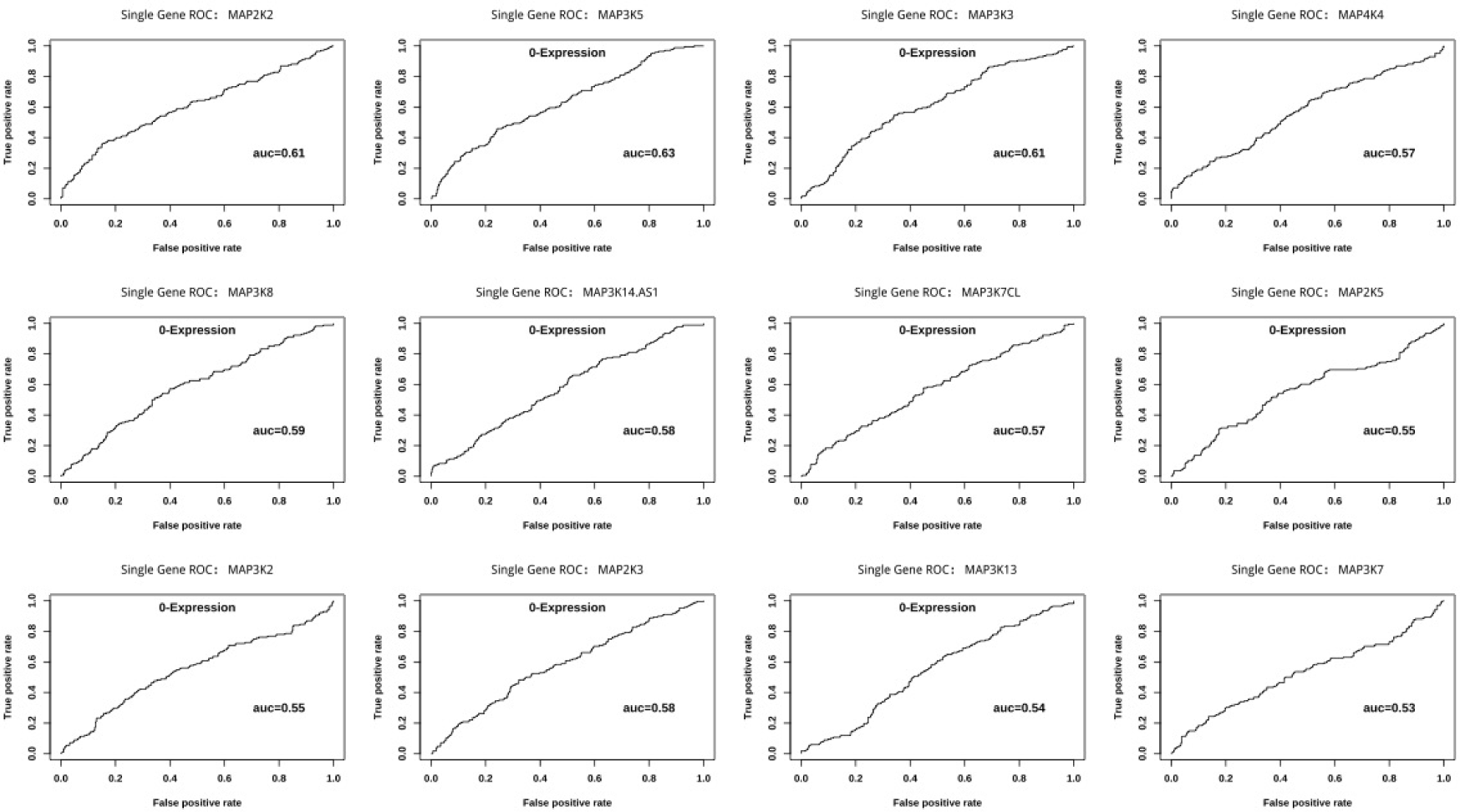
Univariate ROCs of top 12 genes in BRAF cluster in the decreasing order of *P_δ_*.

**Table 4:** AUCs and recurrence risks of 33 BRAF genes ordered by *P_δ_*.

| GENE | AUC | FPR | TPR | $T_g$ | $P_{above}(\%)$ | $P_{below}(\%)$ | $P_\delta(\%)$ | Status |
| --- | --- | --- | --- | --- | --- | --- | --- | --- |
| MAP2K2 | 0.607 | 0.37 | 0.54 | 0.0282 | 44.39 | 27.8 | 16.59 | up |
| MAP3K5 | 0.629 | 0.36 | 0.54 | 0.05 | 26.56 | 40.34 | 13.78 | down |
| MAP3K3 | 0.612 | 0.34 | 0.55 | 0.0439 | 26.74 | 39.35 | 12.61 | down |
| MAP4K4 | 0.568 | 0.51 | 0.64 | -0.0223 | 40.23 | 28.24 | 11.99 | up |
| MAP3K8 | 0.588 | 0.4 | 0.57 | 0.0754 | 27.98 | 39.45 | 11.47 | down |
| MAP3K14-AS1 | 0.576 | 0.51 | 0.64 | -0.0104 | 30.94 | 39.63 | 8.69 | down |
| MAP3K7CL | 0.568 | 0.45 | 0.58 | 0.0071 | 30.22 | 38.91 | 8.69 | down |
| MAP2K5 | 0.552 | 0.39 | 0.54 | 0.0244 | 29.85 | 38.43 | 8.58 | down |
| MAP3K2 | 0.553 | 0.42 | 0.55 | 0.0094 | 30.43 | 38.89 | 8.46 | down |
| MAP2K3 | 0.581 | 0.37 | 0.52 | 0.0306 | 29.73 | 38.05 | 8.32 | down |
| MAP3K13 | 0.541 | 0.48 | 0.58 | -0.0201 | 31.5 | 39.23 | 7.73 | down |
| MAP3K7 | 0.525 | 0.45 | 0.53 | 0.0179 | 38.7 | 31.35 | 7.35 | up |
| MAP3K1 | 0.526 | 0.5 | 0.6 | -0.0183 | 31.7 | 38.71 | 7.01 | down |
| MAP3K6 | 0.543 | 0.46 | 0.57 | 0.0087 | 31.19 | 37.88 | 6.69 | down |
| MAP2K1 | 0.514 | 0.38 | 0.45 | 0.0407 | 38.66 | 32.29 | 6.37 | up |
| RAF1 | 0.536 | 0.39 | 0.46 | 0.0363 | 30.82 | 36.84 | 6.02 | down |
| MAP2K4 | 0.523 | 0.44 | 0.53 | 0.0085 | 31.78 | 37.8 | 6.02 | down |
| MAP3K11 | 0.539 | 0.5 | 0.6 | -0.0042 | 32.07 | 37.55 | 5.48 | down |
| MAP3K9 | 0.542 | 0.5 | 0.6 | -0.0449 | 32.75 | 37.95 | 5.2 | down |
| MAP4K2 | 0.516 | 0.51 | 0.57 | -0.0159 | 37.11 | 32.3 | 4.81 | normal |
| MAP3K21 | 0.525 | 0.47 | 0.53 | 0.0201 | 37.29 | 32.52 | 4.77 | normal |
| MAP4K3 | 0.511 | 0.49 | 0.55 | -0.0021 | 32.5 | 37.19 | 4.69 | normal |
| MAP3K20 | 0.501 | 0.42 | 0.45 | 0.0607 | 32 | 36.48 | 4.48 | normal |
| MAP4K5 | 0.54 | 0.43 | 0.48 | 0.0222 | 32.37 | 36.73 | 4.36 | normal |
| MAP2K6 | 0.506 | 0.36 | 0.4 | 0.0443 | 37.57 | 33.22 | 4.35 | normal |
| BRAF | 0.508 | 0.4 | 0.45 | 0.0458 | 37.31 | 33.1 | 4.21 | normal |
| MAP3K4 | 0.512 | 0.43 | 0.47 | 0.0202 | 37.09 | 33.09 | 4 | normal |
| MAP2K7 | 0.524 | 0.32 | 0.43 | 0.0242 | 32.83 | 36.27 | 3.44 | normal |
| MAP4K1 | 0.514 | 0.5 | 0.55 | -0.0175 | 33.73 | 36.12 | 2.39 | normal |
| MAP3K19 | 0.521 | 0.47 | 0.51 | 0.0101 | 33.64 | 35.88 | 2.24 | normal |
| MAP3K10 | 0.516 | 0.41 | 0.47 | 0.0101 | 33.94 | 35.63 | 1.69 | normal |
| MAP3K14 | 0.519 | 0.37 | 0.48 | 0.059 | 35.75 | 34.32 | 1.43 | normal |
| MAP3K12 | 0.517 | 0.45 | 0.5 | 0.0124 | 35.5 | 34.4 | 1.1 | normal |

#### 3.1.3 EGFR Cluster

EGFR cluster contains 16 members. The ROCs are presented in Figure 3 and the corresponding AUCs, FPRs, TPRs, threshold *T_g_*, population risks are listed in Table 5. There are 12 members with *P_δ_≥* 5%, accounting for 75%, 7 of which including NRG4, EREG, SRC, RGS16, TGFA, CTNNB1 and NRG3 are over-expressed. NRG4, EREG, TGFA and NRG3 are known ligands of EGFR, while RGS16 is phosphorylated by EGFR to have GTPase activation, in addition, EGFR increasingly interacts with SRC and CTNNB1 by phosphorylating MUC1. On the other hand, EGFR itself and MUC1 are down-expressed for lung cancer recurrence, so are other two ligands BTC and AREG. Lastly, The remaining four genes BRAF, NRG2, NRG1 and EGF are normal with *P_δ_ <* 5%.

**Figure 3:**
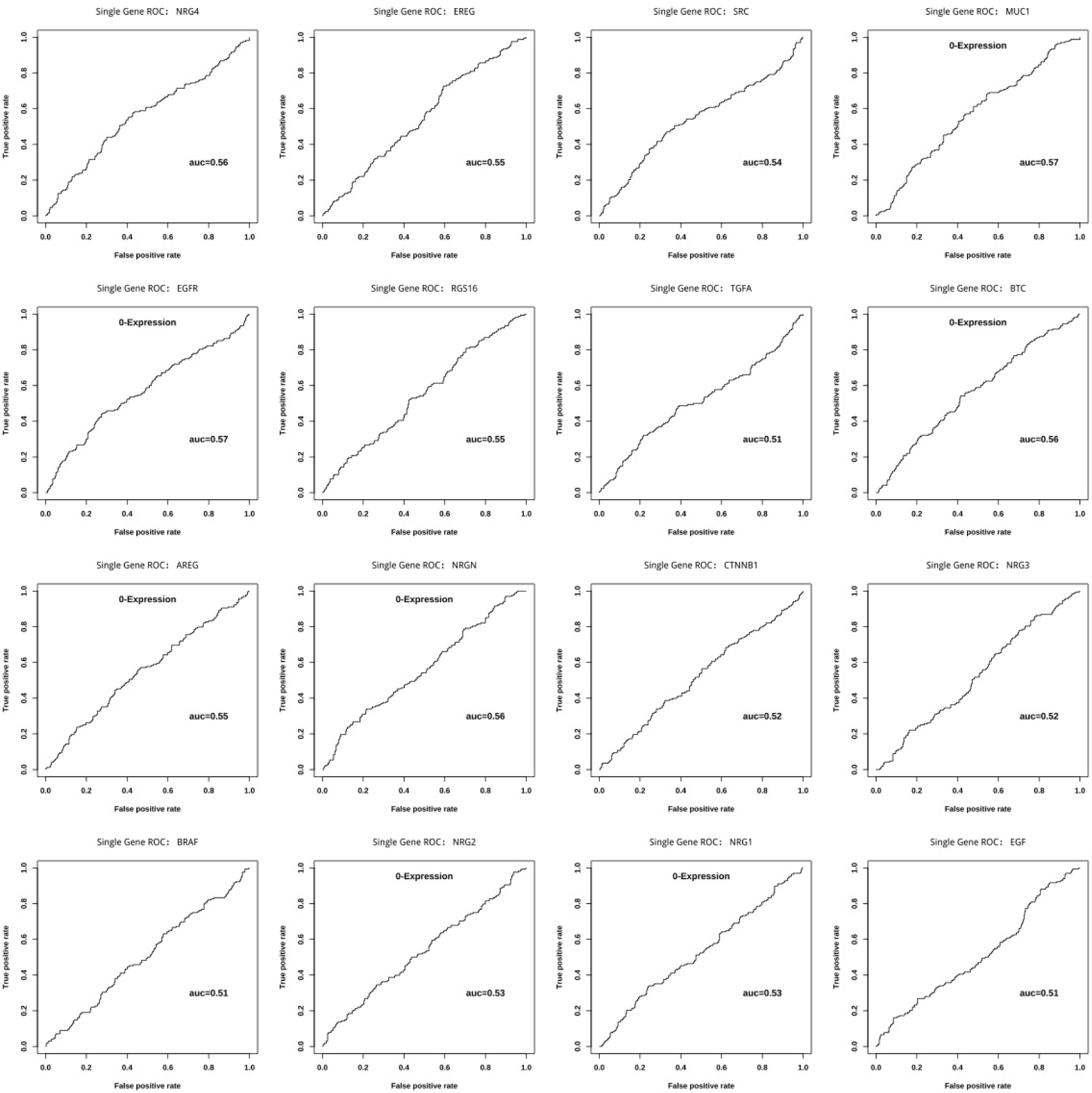
Univariate ROCs of 16 genes in EGFR cluster.

**Table 5:** AUCs and recurrence risks of EGFR genes ordered by *P_δ_*.

| GENE | AUC | FPR | TPR | $T_g$ | $P_{above}(\%)$ | $P_{below}(\%)$ | $P_\delta(\%)$ | Status |
| --- | --- | --- | --- | --- | --- | --- | --- | --- |
| NRG4 | 0.559 | 0.43 | 0.58 | 0.0034 | 41.74 | 28.57 | 13.17 | up |
| EREG | 0.549 | 0.59 | 0.73 | -0.0948 | 39.61 | 26.44 | 13.17 | up |
| SRC | 0.54 | 0.37 | 0.51 | 0.0188 | 42.29 | 29.54 | 12.75 | up |
| MUC1 | 0.567 | 0.43 | 0.56 | 0.0151 | 29.63 | 40.17 | 10.54 | down |
| EGFR | 0.57 | 0.3 | 0.46 | 0.0658 | 29.67 | 38.83 | 9.16 | down |
| RGS16 | 0.549 | 0.43 | 0.52 | 0.0228 | 39.64 | 30.77 | 8.87 | up |
| TGFA | 0.513 | 0.39 | 0.49 | 0.0691 | 39.81 | 31.16 | 8.65 | up |
| BTC | 0.563 | 0.42 | 0.54 | 0.0483 | 30.19 | 38.52 | 8.33 | down |
| AREG | 0.549 | 0.45 | 0.56 | 0.0266 | 30.87 | 38.49 | 7.62 | down |
| NRGN | 0.562 | 0.37 | 0.45 | 0.0989 | 30.81 | 37.1 | 6.29 | down |
| CTNNB1 | 0.52 | 0.5 | 0.57 | -0.0256 | 37.55 | 31.88 | 5.67 | up |
| NRG3 | 0.523 | 0.57 | 0.62 | -0.0287 | 37.1 | 31.66 | 5.44 | up |
| BRAF | 0.508 | 0.4 | 0.45 | 0.0458 | 37.31 | 33.1 | 4.21 | normal |
| NRG2 | 0.532 | 0.44 | 0.5 | 0.0083 | 32.5 | 36.52 | 4.02 | normal |
| NRG1 | 0.528 | 0.4 | 0.45 | 0.0314 | 33.16 | 35.99 | 2.83 | normal |
| EGF | 0.51 | 0.41 | 0.4 | 0.2169 | 34.87 | 34.84 | 0.03 | normal |

#### 3.1.4 MET Cluster

MET cluster contains 8 members. The ROCs are presented in Figure 4 and the corresponding AUCs, FPRs, TPRs, threshold *T_g_*, population risks are listed in Table 6. There are 7 members with *P_δ_ ≥* 5%, accounting for 87.5%. Most MET effectors are PI3-kinase subunits. SRC, GRB2 and PLCG1 are over-expressed while PIK3R1, HGF, GAB1 and MET itself are down-expressed. However, STAT3 is normal with *P_δ_*= 0.09%.

**Figure 4:**
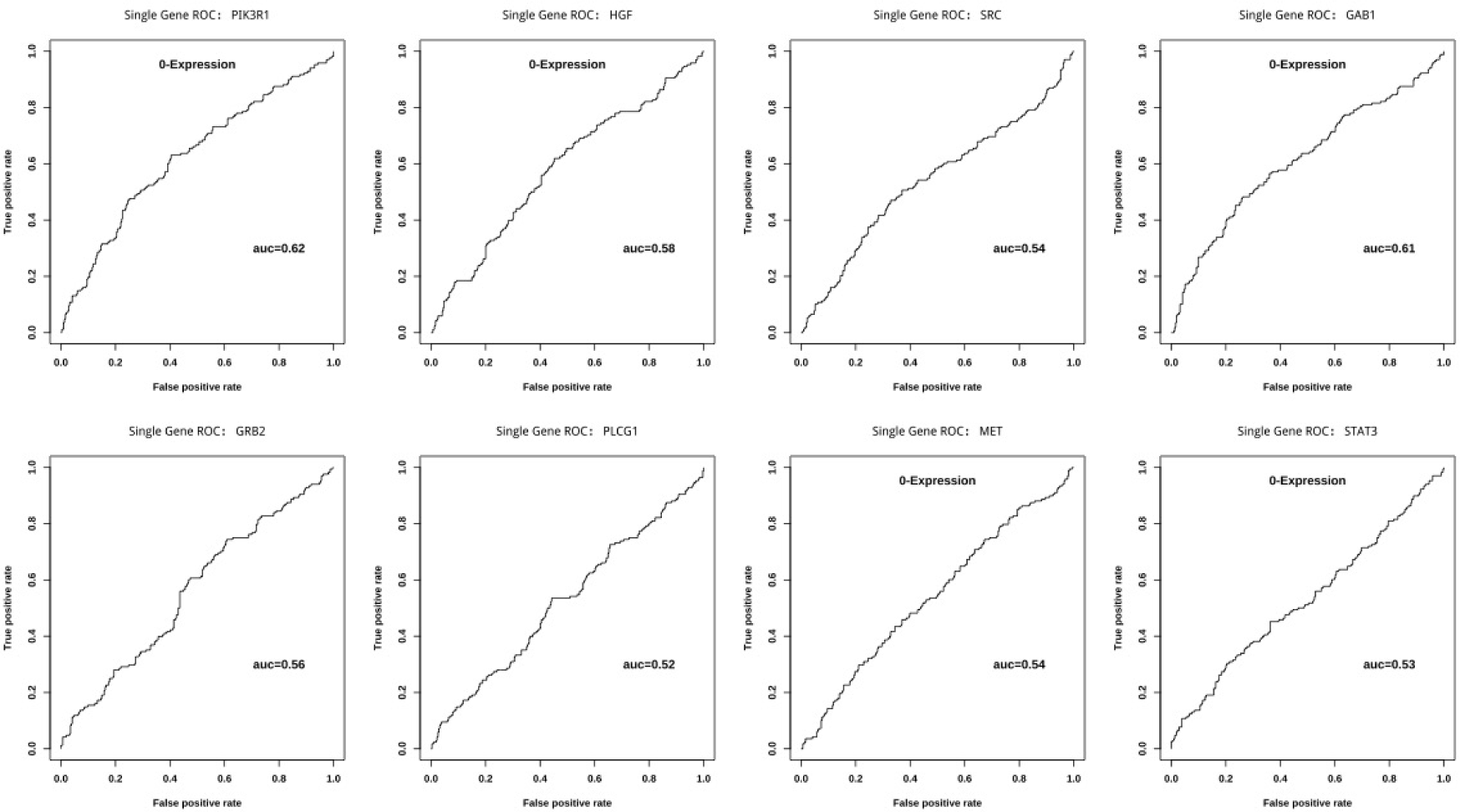
Univariate ROCs of 8 genes in MET cluster.

**Table 6:** AUCs and recurrence risks of MET genes ordered by *P_δ_*.

| GENE | AUC | FPR | TPR | $T_g$ | $P_{above}(\%)$ | $P_{below}(\%)$ | $P_\delta(\%)$ | Status |
| --- | --- | --- | --- | --- | --- | --- | --- | --- |
| PIK3R1 | 0.621 | 0.4 | 0.63 | 0.0369 | 25.76 | 41.2 | 15.44 | down |
| HGF | 0.579 | 0.45 | 0.62 | 0.0079 | 27.23 | 40.89 | 13.66 | down |
| SRC | 0.54 | 0.37 | 0.51 | 0.0188 | 42.29 | 29.54 | 12.75 | up |
| GAB1 | 0.612 | 0.36 | 0.57 | 0.0203 | 27.94 | 39.93 | 11.99 | down |
| GRB2 | 0.559 | 0.47 | 0.6 | -0.0076 | 40.49 | 28.94 | 11.55 | up |
| PLCG1 | 0.524 | 0.44 | 0.54 | 0.0036 | 39.3 | 30.83 | 8.47 | up |
| MET | 0.544 | 0.4 | 0.48 | 0.0288 | 32.23 | 37.5 | 5.27 | down |
| STAT3 | 0.53 | 0.37 | 0.45 | 0.0507 | 34.91 | 34.82 | 0.09 | normal |

#### 3.1.5 NTRK Cluster

NTRK cluster contains 58 members, most of the which are NTRK fusion partners listed in Cocco E et al [12] and are re-organized in Table 7. The ROCs are presented in Figure 5 and the corresponding AUCs, FPRs, TPRs, threshold *T_g_*, population risks are listed in Table 8. There are 37 members with *P_δ_≥* 5%, accounting for 64%, within which 24 are over-expressed: ETV6, TPM3, SLITRK1, TFG, SLITRK4, CHTOP, SLITRK5, TPM4, TP53, TRAF2, AGBL3, LYN, RFWD2, NTRK1, AFAP1, AGBL5, UBE2R2, SQSTM1, SLITRK2, MRPL24, NTRK2, GRIPAP1, SLITRK6 and TRIM24 and 13 are down-expressed: MPRIP, TLE4, RBPMS, NFASC, NTRK3, ARHGEF2, CD74, RABGAP1L, NACC2, TRIM63, IGFBP7, DAB2IP, AGBL1. The remaining 24 genes: AGBL2, PPL, BCR, SCYL3, LMNA, MYO5A, CTRC, PLEKHA6, BCAN, PDE4DIP, HNRNPA2B1, VCL, TPR, PAN3, QKI, SLITRK3, EML4, BTBD1, STRN, LRRC71 and IRF2BP2 don’t show much differences with *P_δ_ <* 5%.

**Figure 5:**
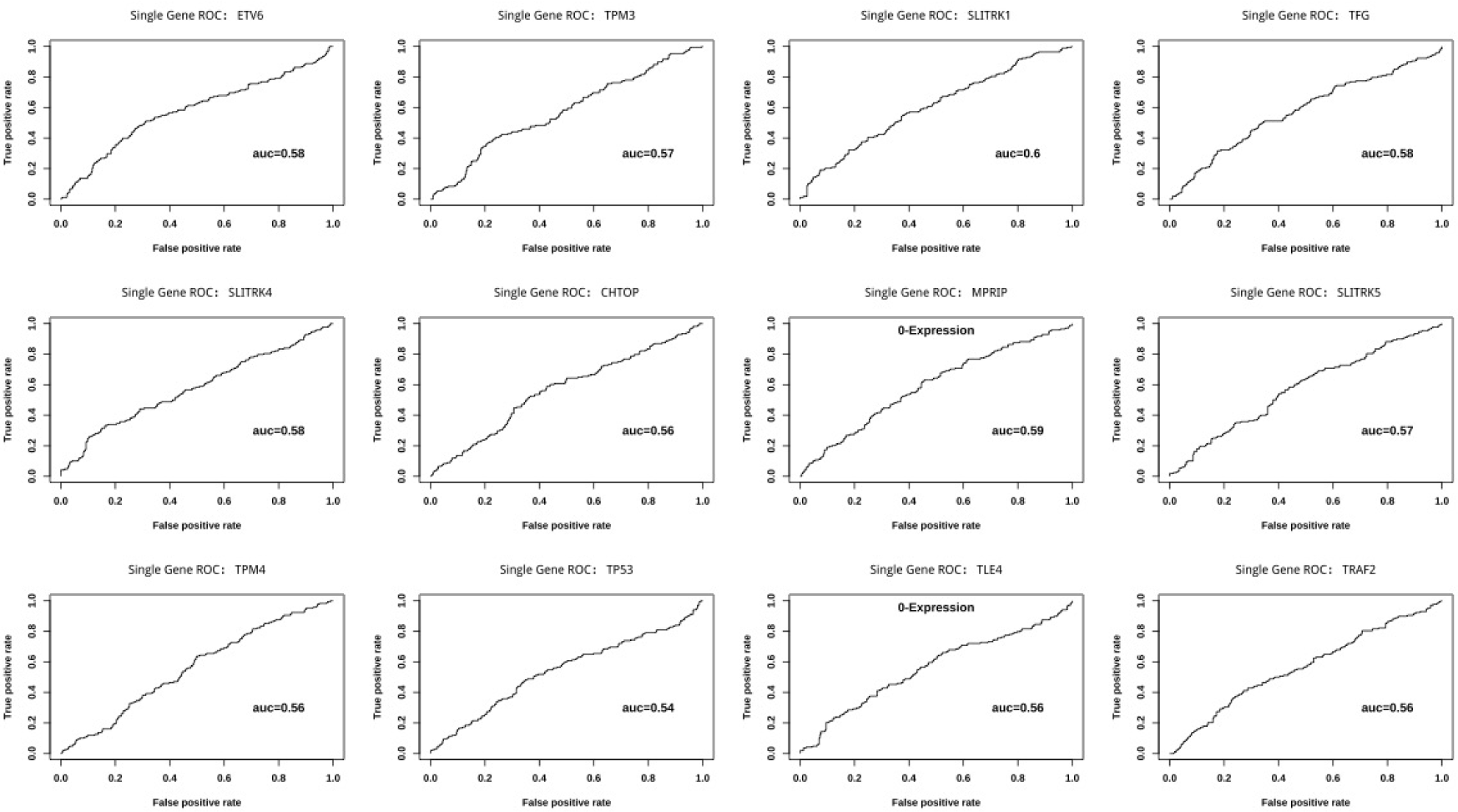
Univariate ROCs of top 12 genes in NTRK cluster.

**Table 7:**
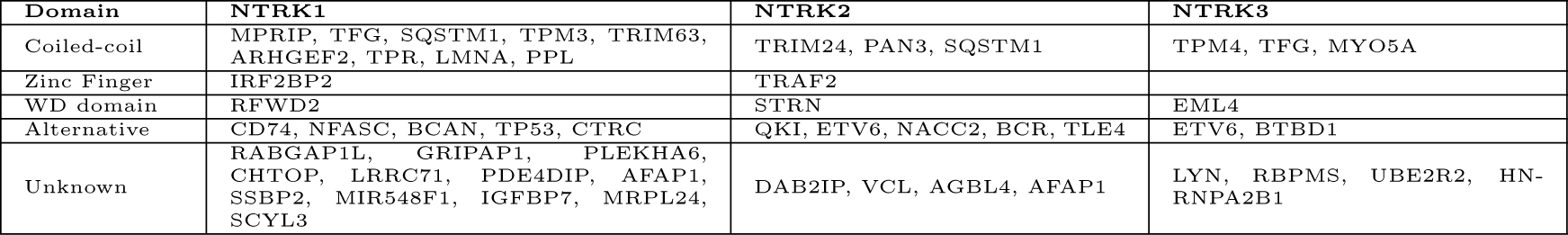
NTRK fusion partners as listed in Cocco E et al [12]

**Table 8:**
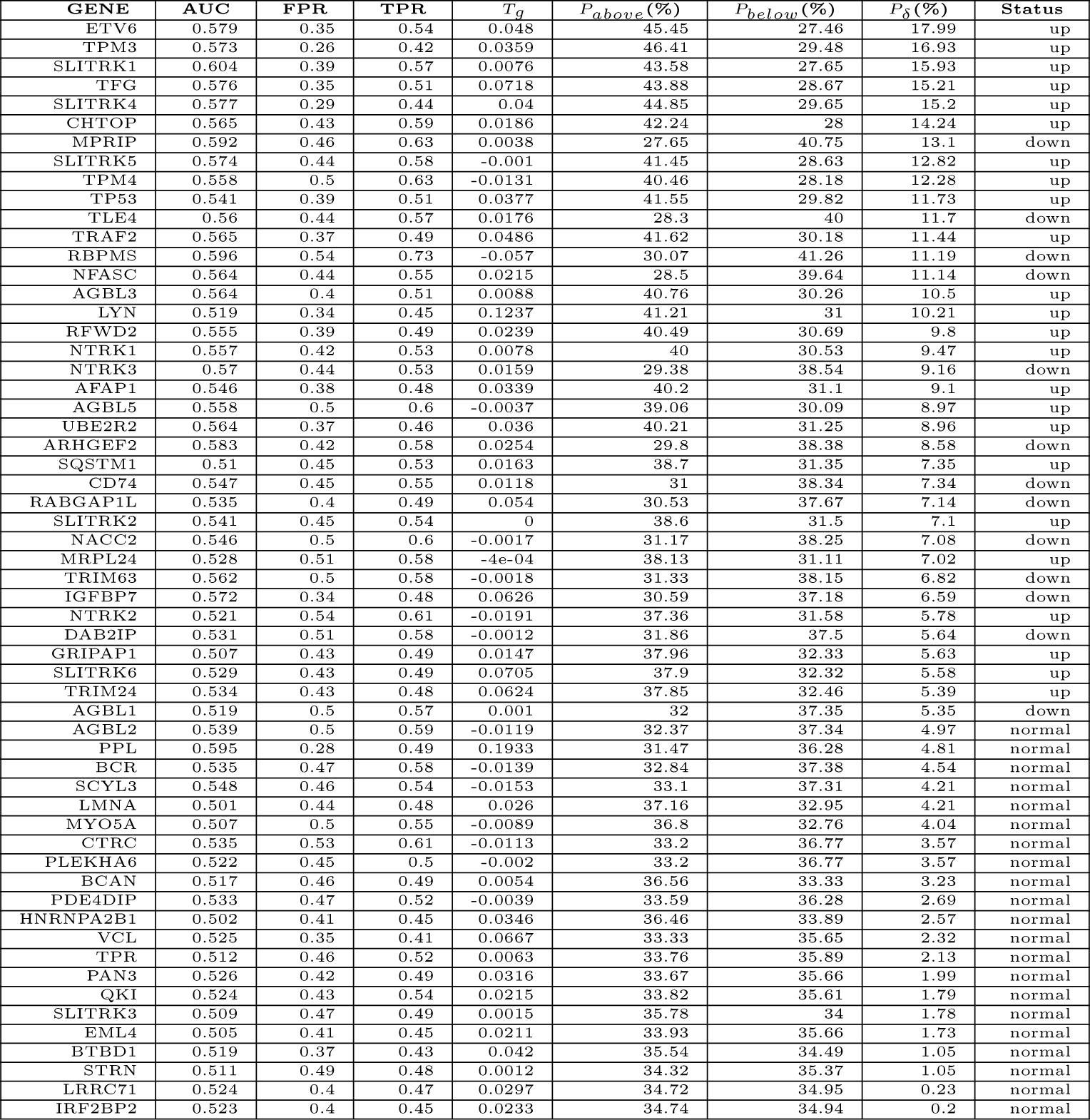
AUCs and recurrence risks of NTRK genes ordered by *P_δ_*.

| GENE | AUC | FPR | TPR | $T_g$ | $P_{above}(\%)$ | $P_{below}(\%)$ | $P_\delta(\%)$ | Status |
| --- | --- | --- | --- | --- | --- | --- | --- | --- |
| ETV6 | 0.579 | 0.35 | 0.54 | 0.048 | 45.45 | 27.46 | 17.99 | up |
| TPM3 | 0.573 | 0.26 | 0.42 | 0.0359 | 46.41 | 29.48 | 16.93 | up |
| SLITRK1 | 0.604 | 0.39 | 0.57 | 0.0076 | 43.58 | 27.65 | 15.93 | up |
| TFG | 0.576 | 0.35 | 0.51 | 0.0718 | 43.88 | 28.67 | 15.21 | up |
| SLITRK4 | 0.577 | 0.29 | 0.44 | 0.04 | 44.85 | 29.65 | 15.2 | up |
| CHTOP | 0.565 | 0.43 | 0.59 | 0.0186 | 42.24 | 28 | 14.24 | up |
| MPRIP | 0.592 | 0.46 | 0.63 | 0.0038 | 27.65 | 40.75 | 13.1 | down |
| SLITRK5 | 0.574 | 0.44 | 0.58 | -0.001 | 41.45 | 28.63 | 12.82 | up |
| TPM4 | 0.558 | 0.5 | 0.63 | -0.0131 | 40.46 | 28.18 | 12.28 | up |
| TP53 | 0.541 | 0.39 | 0.51 | 0.0377 | 41.55 | 29.82 | 11.73 | up |
| TLE4 | 0.56 | 0.44 | 0.57 | 0.0176 | 28.3 | 40 | 11.7 | down |
| TRAF2 | 0.565 | 0.37 | 0.49 | 0.0486 | 41.62 | 30.18 | 11.44 | up |
| RBPMS | 0.596 | 0.54 | 0.73 | -0.057 | 30.07 | 41.26 | 11.19 | down |
| NFASC | 0.564 | 0.44 | 0.55 | 0.0215 | 28.5 | 39.64 | 11.14 | down |
| AGBL3 | 0.564 | 0.4 | 0.51 | 0.0088 | 40.76 | 30.26 | 10.5 | up |
| LYN | 0.519 | 0.34 | 0.45 | 0.1237 | 41.21 | 31 | 10.21 | up |
| RFWD2 | 0.555 | 0.39 | 0.49 | 0.0239 | 40.49 | 30.69 | 9.8 | up |
| NTRK1 | 0.557 | 0.42 | 0.53 | 0.0078 | 40 | 30.53 | 9.47 | up |
| NTRK3 | 0.57 | 0.44 | 0.53 | 0.0159 | 29.38 | 38.54 | 9.16 | down |
| AFAP1 | 0.546 | 0.38 | 0.48 | 0.0339 | 40.2 | 31.1 | 9.1 | up |
| AGBL5 | 0.558 | 0.5 | 0.6 | -0.0037 | 39.06 | 30.09 | 8.97 | up |
| UBE2R2 | 0.564 | 0.37 | 0.46 | 0.036 | 40.21 | 31.25 | 8.96 | up |
| ARHGEF2 | 0.583 | 0.42 | 0.58 | 0.0254 | 29.8 | 38.38 | 8.58 | down |
| SQSTM1 | 0.51 | 0.45 | 0.53 | 0.0163 | 38.7 | 31.35 | 7.35 | up |
| CD74 | 0.547 | 0.45 | 0.55 | 0.0118 | 31 | 38.34 | 7.34 | down |
| RABGAP1L | 0.535 | 0.4 | 0.49 | 0.054 | 30.53 | 37.67 | 7.14 | down |
| SLITRK2 | 0.541 | 0.45 | 0.54 | 0 | 38.6 | 31.5 | 7.1 | up |
| NACC2 | 0.546 | 0.5 | 0.6 | -0.0017 | 31.17 | 38.25 | 7.08 | down |
| MRPL24 | 0.528 | 0.51 | 0.58 | -4e-04 | 38.13 | 31.11 | 7.02 | up |
| TRIM63 | 0.562 | 0.5 | 0.58 | -0.0018 | 31.33 | 38.15 | 6.82 | down |
| IGFBP7 | 0.572 | 0.34 | 0.48 | 0.0626 | 30.59 | 37.18 | 6.59 | down |
| NTRK2 | 0.521 | 0.54 | 0.61 | -0.0191 | 37.36 | 31.58 | 5.78 | up |
| DAB2IP | 0.531 | 0.51 | 0.58 | -0.0012 | 31.86 | 37.5 | 5.64 | down |
| GRIPAP1 | 0.507 | 0.43 | 0.49 | 0.0147 | 37.96 | 32.33 | 5.63 | up |
| SLITRK6 | 0.529 | 0.43 | 0.49 | 0.0705 | 37.9 | 32.32 | 5.58 | up |
| TRIM24 | 0.534 | 0.43 | 0.48 | 0.0624 | 37.85 | 32.46 | 5.39 | up |
| AGBL1 | 0.519 | 0.5 | 0.57 | 0.001 | 32 | 37.35 | 5.35 | down |
| AGBL2 | 0.539 | 0.5 | 0.59 | -0.0119 | 32.37 | 37.34 | 4.97 | normal |
| PPL | 0.595 | 0.28 | 0.49 | 0.1933 | 31.47 | 36.28 | 4.81 | normal |
| BCR | 0.535 | 0.47 | 0.58 | -0.0139 | 32.84 | 37.38 | 4.54 | normal |
| SCYL3 | 0.548 | 0.46 | 0.54 | -0.0153 | 33.1 | 37.31 | 4.21 | normal |
| LMNA | 0.501 | 0.44 | 0.48 | 0.026 | 37.16 | 32.95 | 4.21 | normal |
| MYO5A | 0.507 | 0.5 | 0.55 | -0.0089 | 36.8 | 32.76 | 4.04 | normal |
| CTRC | 0.535 | 0.53 | 0.61 | -0.0113 | 33.2 | 36.77 | 3.57 | normal |
| PLEKHA6 | 0.522 | 0.45 | 0.5 | -0.002 | 33.2 | 36.77 | 3.57 | normal |
| BCAN | 0.517 | 0.46 | 0.49 | 0.0054 | 36.56 | 33.33 | 3.23 | normal |
| PDE4DIP | 0.533 | 0.47 | 0.52 | -0.0039 | 33.59 | 36.28 | 2.69 | normal |
| HNRNPA2B1 | 0.502 | 0.41 | 0.45 | 0.0346 | 36.46 | 33.89 | 2.57 | normal |
| VCL | 0.525 | 0.35 | 0.41 | 0.0667 | 33.33 | 35.65 | 2.32 | normal |
| TPR | 0.512 | 0.46 | 0.52 | 0.0063 | 33.76 | 35.89 | 2.13 | normal |
| PAN3 | 0.526 | 0.42 | 0.49 | 0.0316 | 33.67 | 35.66 | 1.99 | normal |
| QKI | 0.524 | 0.43 | 0.54 | 0.0215 | 33.82 | 35.61 | 1.79 | normal |
| SLITRK3 | 0.509 | 0.47 | 0.49 | 0.0015 | 35.78 | 34 | 1.78 | normal |
| EML4 | 0.505 | 0.41 | 0.45 | 0.0211 | 33.93 | 35.66 | 1.73 | normal |
| BTBD1 | 0.519 | 0.37 | 0.43 | 0.042 | 35.54 | 34.49 | 1.05 | normal |
| STRN | 0.511 | 0.49 | 0.48 | 0.0012 | 34.32 | 35.37 | 1.05 | normal |
| LRRC71 | 0.524 | 0.4 | 0.47 | 0.0297 | 34.72 | 34.95 | 0.23 | normal |
| IRF2BP2 | 0.523 | 0.4 | 0.45 | 0.0233 | 34.74 | 34.94 | 0.2 | normal |

As shown in Table 7, in the first row ETV6 is over-expressed with *P_above_* = 45.45%, *P_below_* = 27.46% and a difference of *P_δ_* = 17.99%. ETV6 is an ETS family transcription factor repressing transcription. ETV6-NTRK3 fusion has been found in different types of cancers and its expression activates the MAPK and PI3K pathways [12]. The second is actin-binding TPM3, tropomyosin 3, with *P_δ_*= 16.93%. TPM3-NTRK1 fusion has been reported broadly in many different tumor types, but it is very rare in lung cancer. TPM3-NTRK1 fusion was confirmed in a Chinese lung cancer study [74]. Choi et al. [14] reported a NSCLC case of acquired TPM3-NTRK1 fusion resistant to larotrectinib with EML4-ALK fusion progressed on lorlatinib. For NTRK family itself, NTRK1/3 having *P_δ_* around 9% and NTRK2 having *P_δ_* = 5.78%, however NTRK1/2 are over-expressed while NTRK3 is down-expressed. Additionally, all 6 SLIT and NTRK like family members [6] were selected into the NTRK cluster and they are overexpressed. SLITTRK1/4/5 have *P_δ_* greater than 12%, SLITTRK2/6 have modest *P_δ_*around7% while SLITTRK3 has neglective *P_δ_* = 1.78%. SLITRK5 mediates BDNF-dependent NTRK2 (TrkB) trafficking and signaling [56]. SLITRK3 activates NTRK3 in squamous cell lung cancer [8]. On the other hand, MPRIP stands at the top with *P_δ_* = 13.1%. MPRIP-NTRK1 and CD74-NTRK1 fusions were identified by Vaishnavi A et al. [61] while CD74 has modest *P_δ_*= 7.34%, Both fusions lead to TRKA kinase activity. MPRIP targets myosin phosphatase to the actin cytoskeleton and enables cadherin binding. MPRIP can be also fusion partner of other drive gene in lung cancer. A lung cancer case was reported to be sensitive to ALK inhibitor with MPRIP-ALK fusion[19]. Another late stage case was shown to be sensitive to crizotinib with MPRIP-ROS1 fusion [54]. The second on the down-regulation side is transcription corepressor TLE4, which inhibits the transcriptional activation mediated by PAX5, by CTNNB1 and by TCF family members in Wnt signaling.

#### 3.1.6 RAS Cluster

RAS cluster contains 35 members. The ROCs are presented in Figure 6 and the corresponding AUCs, FPRs, TPRs, threshold *T_g_*, population risks are listed in Table 9. There are 25 members with *P_δ_≥* 5% accounting for 71%, with 13 over-expressed and 12 down-expressed, and the remaining 12 are normal with *P_δ_ <* 5%. They functionally belong to the following categories:

**Figure 6:**
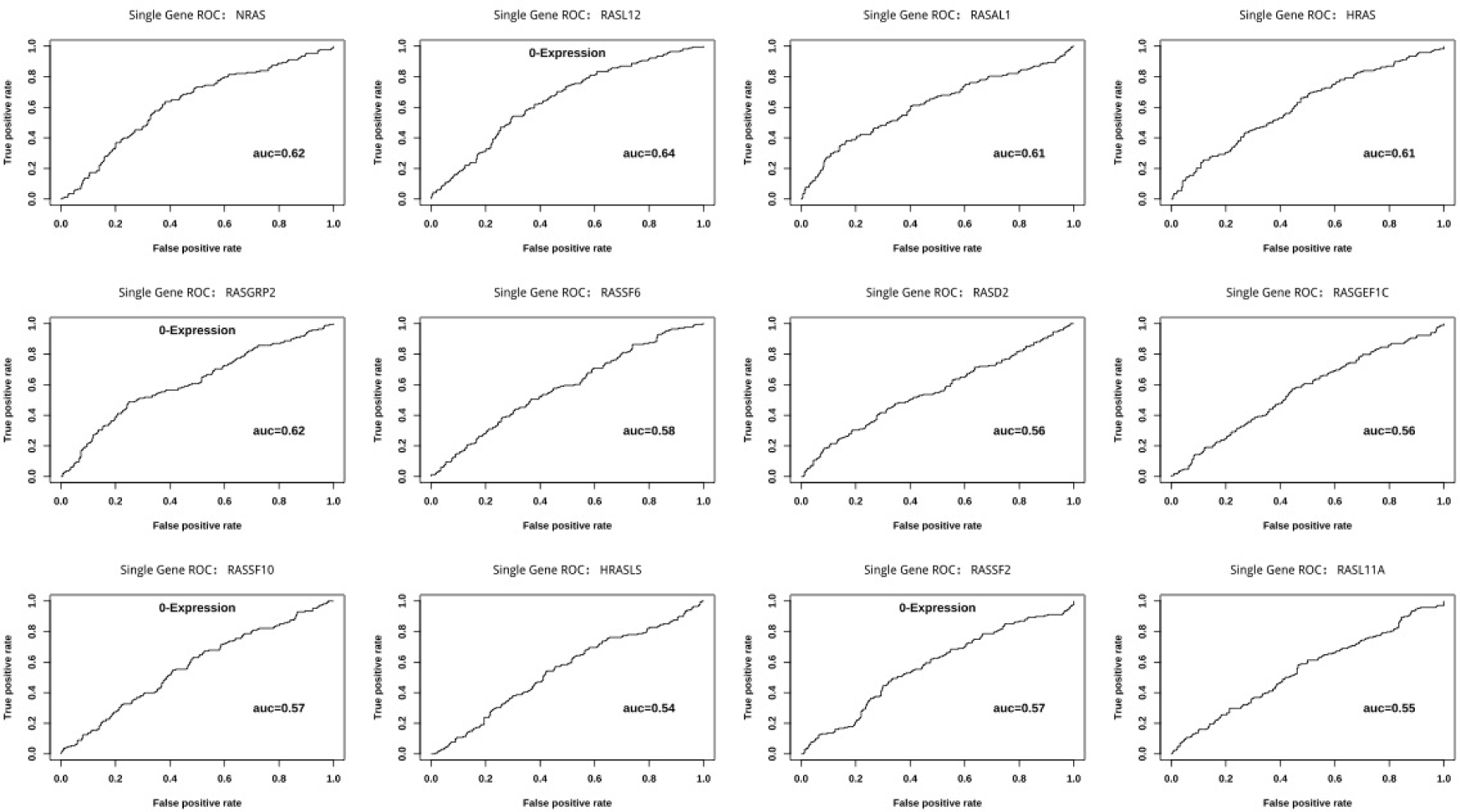
Univariate ROCs of top 12 genes in RAS cluster.

**Table 9:**
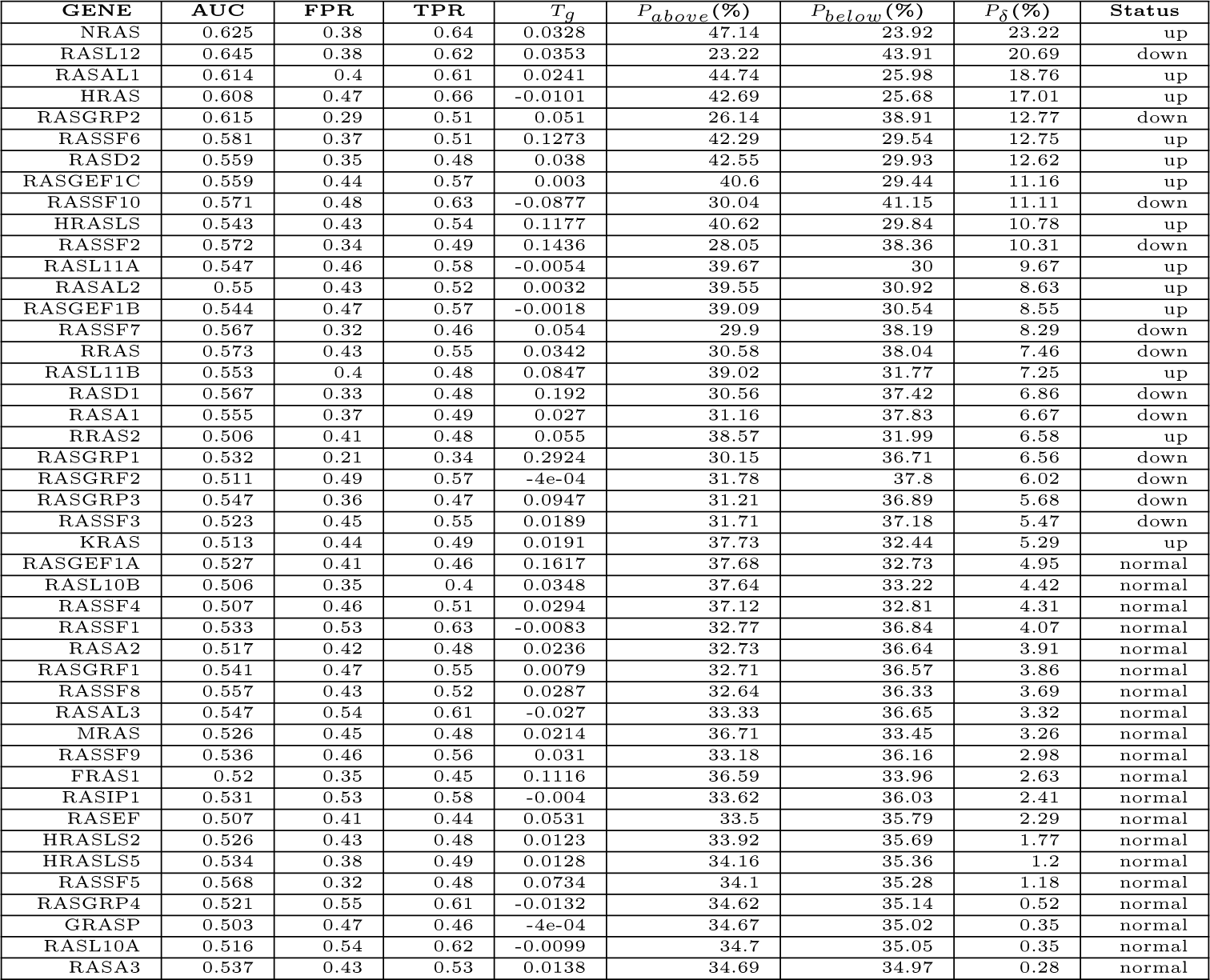
AUCs and recurrence risks of RAS genes ordered by *P_δ_*.

- *Ras/Rab GTPases* over-expressed HRAS, NRAS, KRAS, RRAS2, RASD2; down-expressed RASD1, RRAS; normal MRAS.
- *RAS like family or HRAS like suppressors* over-expressed RASL11A/11B, HRAS like suppressor HRASLS; down-expressed RASL12; and normal RASL10A/10B, HRASLS2/5.
- *Ras-association domain family (Rassf)* over-expressed RASSF6; down-expressed RASSF2/3/7/10; and normal RASSF1/4/5/8/9.
- *RasGAP (GTPase activating protein)* over-expressed RASAL1/2; down-expressed RASA1; normal RASAL3, RASA2/3. RASAL1 belongs to GAP1 and suppresses RAS function. RASAL2 encodes a characteristic domain of GAP and inhibits Ras-cyclic AMP pathway. RASAL3 encodes a protein with pleckstrin homology (PH), C2, and RasGAP domains and is important for liver natural killer T (NKT) cell expansion and functions by suppressing RAS activity and the down-stream ERK signaling pathway.
- *RasGEF (guanine nucleotide exchange factor)* over-expressed RASGEF1B/1C and normal RASGEF1A. RASGEF1A is specific for RAP2A, KRAS, HRAS, and NRAS in vivo. RASGEF1B is only specific for RAP2A. Moreover, it also includes guanyl-releasing factors (GRF), RASGRF2, down-expressed and normal RASGRF1; and guanyl-releasing proteins (GRP), down-expressed RASGRP1/2/3, and normal RASGRP4.

The top tier of *P_δ_*in between 17% and 25% contains 4 genes, in which NRAS, HRAS and RASAL1 are up and RASL12 is down. KRAS is also up but with modest *P_δ_*= 5.29%. This is in consistent with the finding that lung cancer patients with lower RAS expression and treated with bevacizumab plus chemotherapy had a longer PFS and OS than with high RAS expression[5]. Interestingly, NRAS and HRAS suppress KRAS-driven lung cancer growth[60].

#### 3.1.7 RET Cluster

RET cluster contains 72 members, most of which are fusion partners [42]. The ROCs are presented in Figure 7 and the corresponding AUCs, FPRs, TPRs, threshold *T_g_*, population risks are listed in Table 10. There are 47 members with *P_δ_ ≥* 5%, accounting for 65%, within which 18 over-expressed and 29 down-expressed. The rest 25 members are normal. The over-expressed ones include MRPS30, CDC123, LSM14A, IL2RA, GPRC5B, KIAA1217, UBE2D1, PRPF18, PARD3, RETNLB, CLIP1, GFRA3, RET, KIAA1468, TRIM33, GDNF, TRIM24, RETREG1; The down ones include ANK3, GFRA1, EPC1, CCDC186, MPRIP, NCOA4, RETN, SORBS1, MINDY3, PRKAR1A, DOCK1, RBPMS, KIF13A, SIRT1, ARHGAP12, MYO5C, ZNF438, WAC, RETSAT, KIF5B, CCDC88C, TSSK4, CCDC3, PCM1, TBC1D32, PRKCQ, NRP1, PRKG1, PICALM; The normal ones are: CTNNA3, GFRA2, PTPRK, PTK2, RASSF4, DYDC1, CUX1, RUFY2, EPHA5, ADD3, ANKS1B, CCNY, DUSP5, FRMD4A, PTER, ZNF43, GFRA4, RETREG3, EML4, ERC1, CCNYL1, EML6, RETREG2, CCDC6, ALOX5. Among the top 21 genes with *P_δ_≥* 8%, there are only 5 over-expressed ones and the rest 16 are down ones.

**Figure 7:**
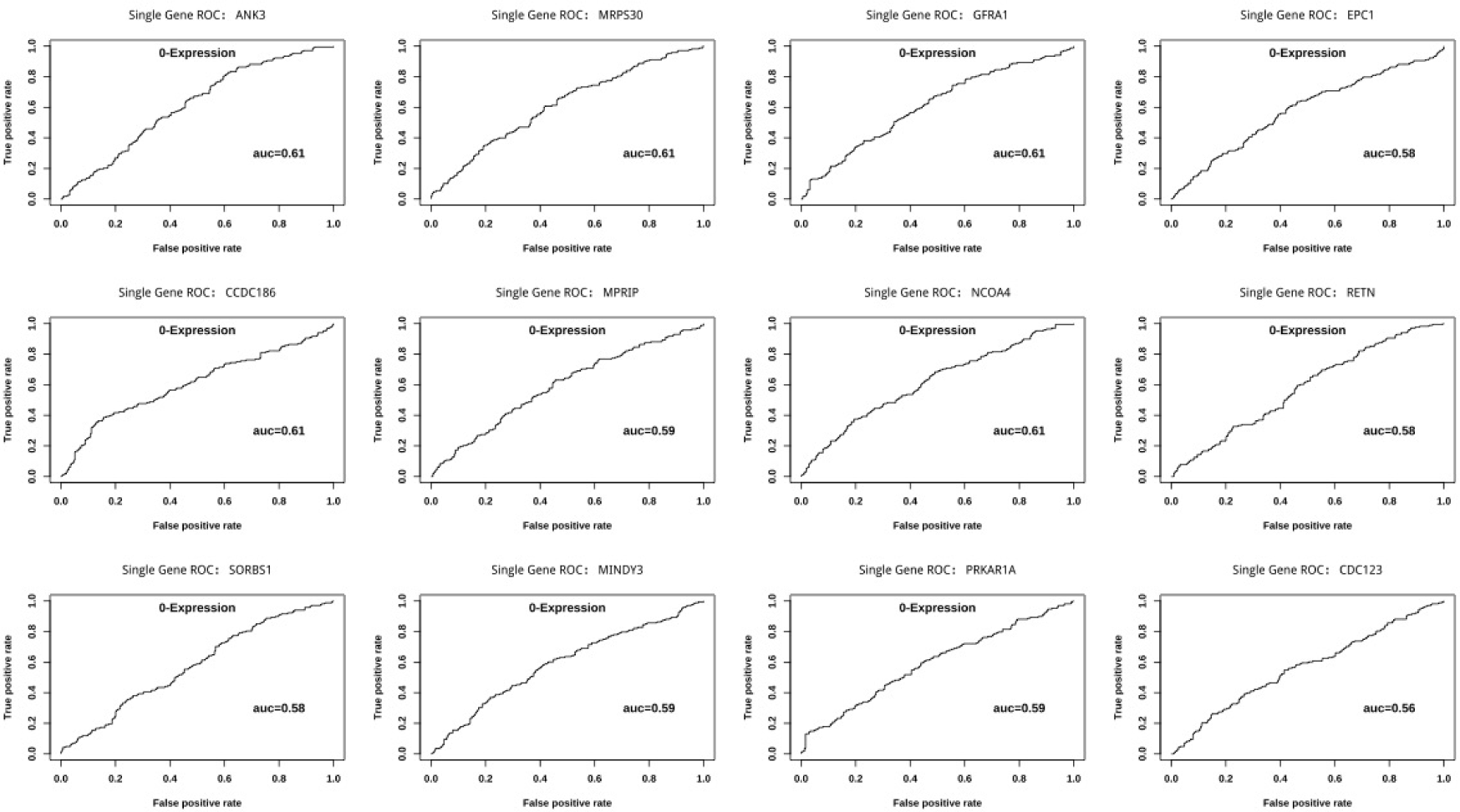
Univariate ROCs of top 12 genes in RET cluster.

**Table 10:**
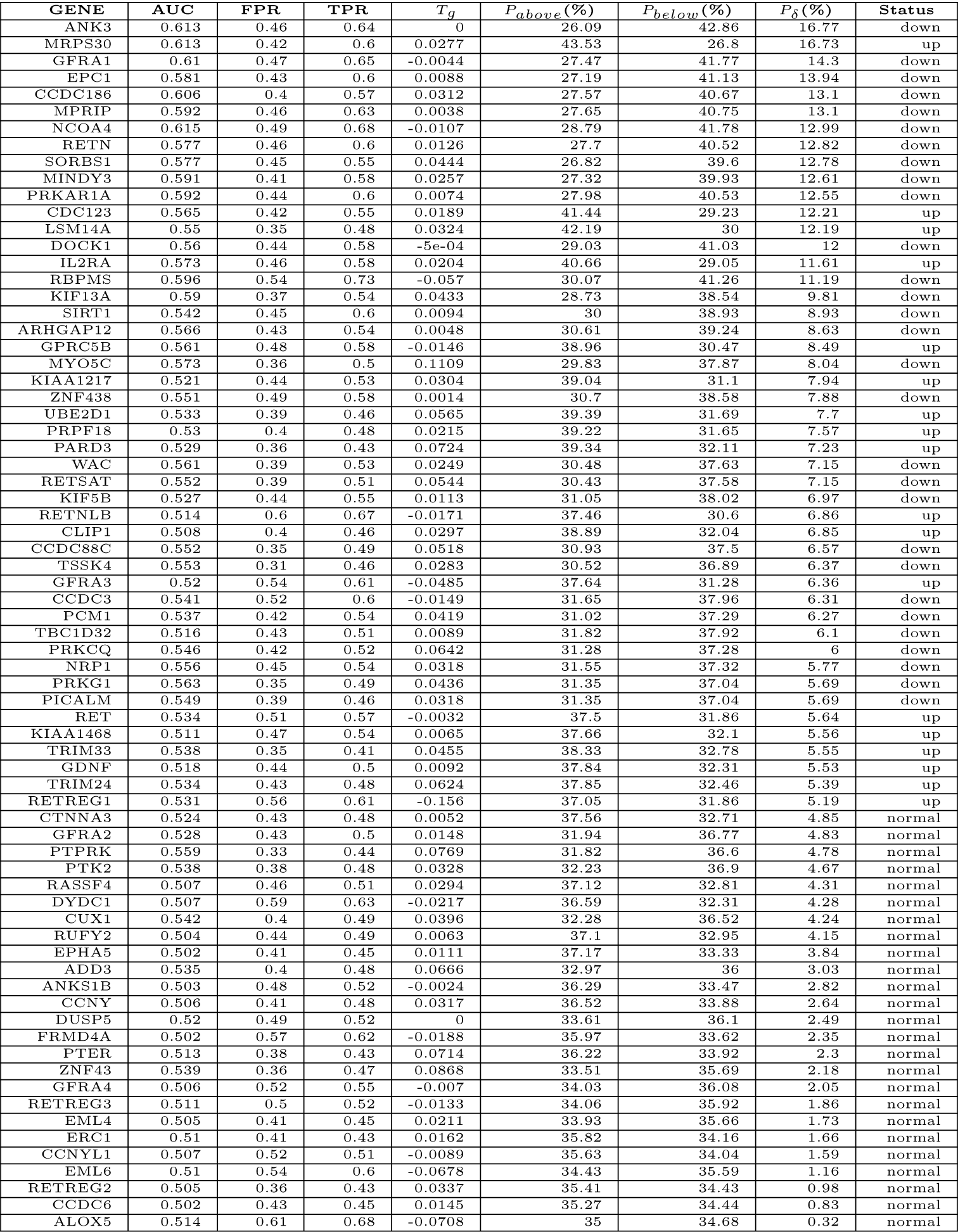
AUCs and recurrence risks of RET genes ordered by *P_δ_*.

#### 3.1.8 ROS1 Cluster

ROS1 cluster contains 33 members, most of which are fusion partners [43]. The ROCs are presented in Figure 8 and the corresponding AUCs, FPRs, TPRs, threshold *T_g_*, population risks are listed in Table 11. There are 21 members with *P_δ_≥* 5%, accounting for 64%, within which PTPN11, TPM3, TFG, KDELR2, CEP72, TPD52L1, VAV3, CLTC, WNK1 are over-expressed, and SLC34A2, SDC4, RBPMS, LRIG3, SLMAP, KMT2C, PLCG2, MYO5C, PROS1, CD74, EZR, ROS1 are down-expressed, and the rest 12 genes MSN, MAPK1, TMEM106B, SLC6A17, MAPK3, LIMA1, ZCCHC8, IRS1, GOPC, CCDC6, AKT1, STAT3 are normal with *P_δ_ <* 5%. At the top, PTPN11, named as protein tyrosine phosphatase non-receptor type 11, more commonly aliased as SHP2, has the highest *P_δ_* = 20.71% and is over-expressed for higher risk recurrence. ROS1 mediates the phosphorylation of PTPN11 to activate the downstream pathway. The second top over-expressed is actin-binding TPM3 with *P_δ_*= 16.93%, which also appears in NTRK cluster. A case reported that EML4-ALK and TPM3-ROS1 fusion coexistence in an advanced NSCLC Chinese man[76]. The third over-expressed is TFG with *P_δ_* = 15.21%, called trafficking from ER to Golgi regulator, also called TRK-fused gene protein, is required for secretory cargo traffic from the ER to the Golgi apparatus, TFG-ROS1 fusion was reported in lung cancers[1] and other cancers[4]. On the down-expression side, SLC34A2 is at the top with *P_δ_*= 11.76%. SLC34A2-ROS1 fusion was reported in lung cancer tissues[16]. SDC4, RBPMS and LRIG3 are the next 3 down-expressed genes and with similar *P_δ_*. SDC4 is a cell surface proteoglycan that bears heparan sulfate. SDC4-ROS1 fusion is rare in lung cancer, a case was reported that SDC4-ROS1 fusion positive was treated with crizotinib followed by three cycles of chemotherapy, after disease progression it was revealed the original SDC4-ROS1 fusion along with a KRAS point mutation (p.G12D)[78].

**Figure 8:**
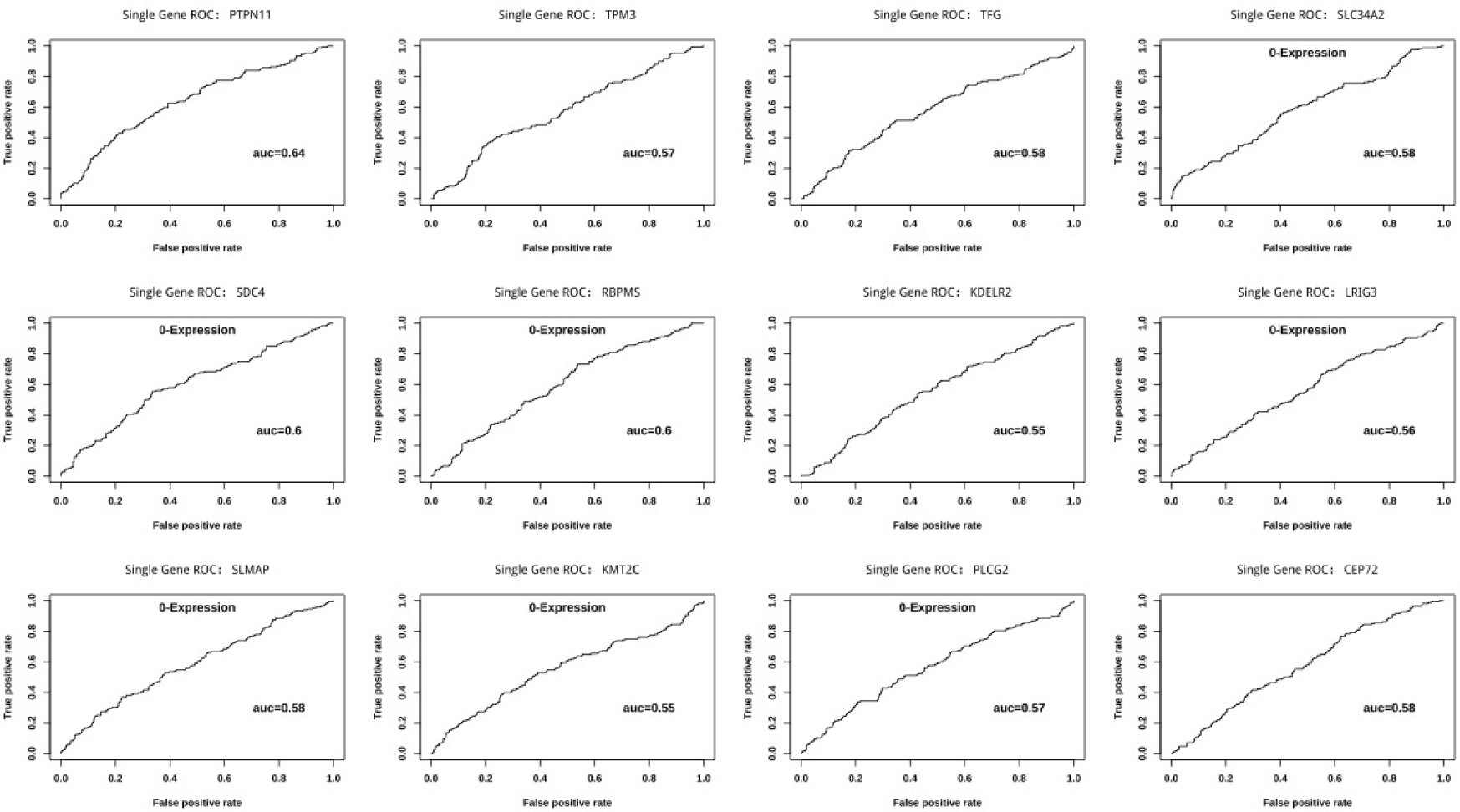
Univariate ROCs of top 12 genes in ROS1 cluster.

**Table 11:**
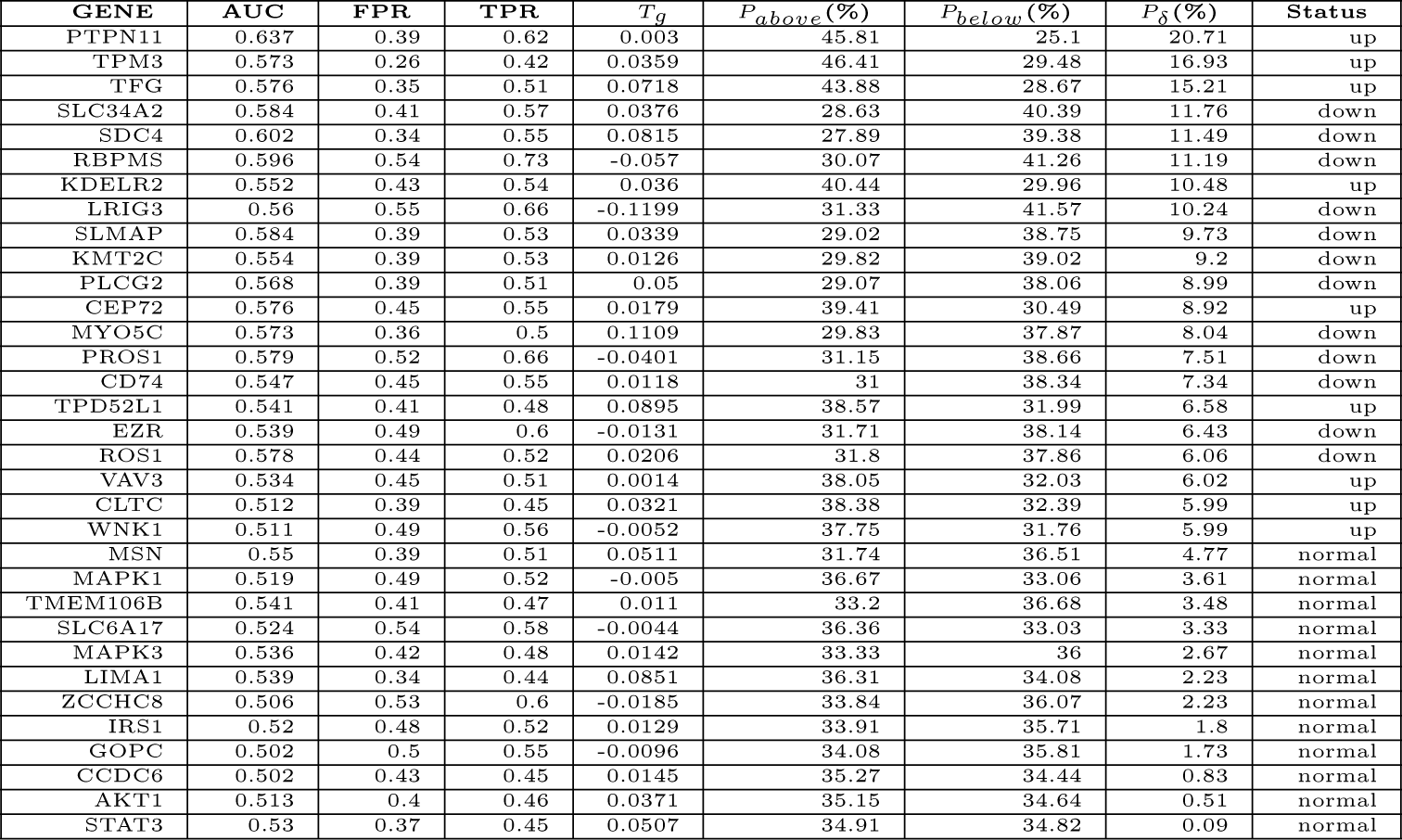
AUCs and recurrence risks of ROS1 genes ordered by *P_δ_*.

#### 3.1.9 TP53 Cluster

TP53 cluster contains 11 members. The ROCs are presented in Figure 9 and the corresponding AUCs, FPRs, TPRs, threshold *T_g_*, population risks are listed in Table 12. There are 9 members with *P_δ_ ≥* 5% within which TP53, TP53BP1, TP53BP2, TP53I13, TP53I3, TP53INP2, TP53RK are over-expressed while TP53INP1, TP53TG5 are down. The rest two normal ones are TP53I11 and TP53TG1. At the top is down-expressed TP53INP1 with the highest *P_δ_*= 18.42% while the second is the over-expressed TP53BP2 with *P_δ_* = 16%. Unlike other clusters of which the seeds have modest *P_δ_*, TP53 itself is over-expressed and stands at the third with *P_δ_* = 11.73%. TP53INP1, named as tumor protein p53-inducible nuclear protein 1, is a tumor suppressor, over-expressed during stress responses including inflammation and regulating metabolic homeostasis[50]. Moreover it plays important role in DNA damage response[52]. On the contrary, TP53INP2 is over-expressed with notable *P_δ_* = 9.83% and it plays dual roles and switches between transcription and autophagy by sensing the nutrient status [70]. TP53BP2, P53-binding protein 2, also called apoptosis stimulating protein 2 of P53 (ASPP2), is involved with multiple pathways in tumorigenesis[20]. Similarly, TP53BP1 is also over-expressed but with modest *P_δ_* = 7.94% and plays critical roles in DNA damage response in cancer[36].

**Figure 9:**
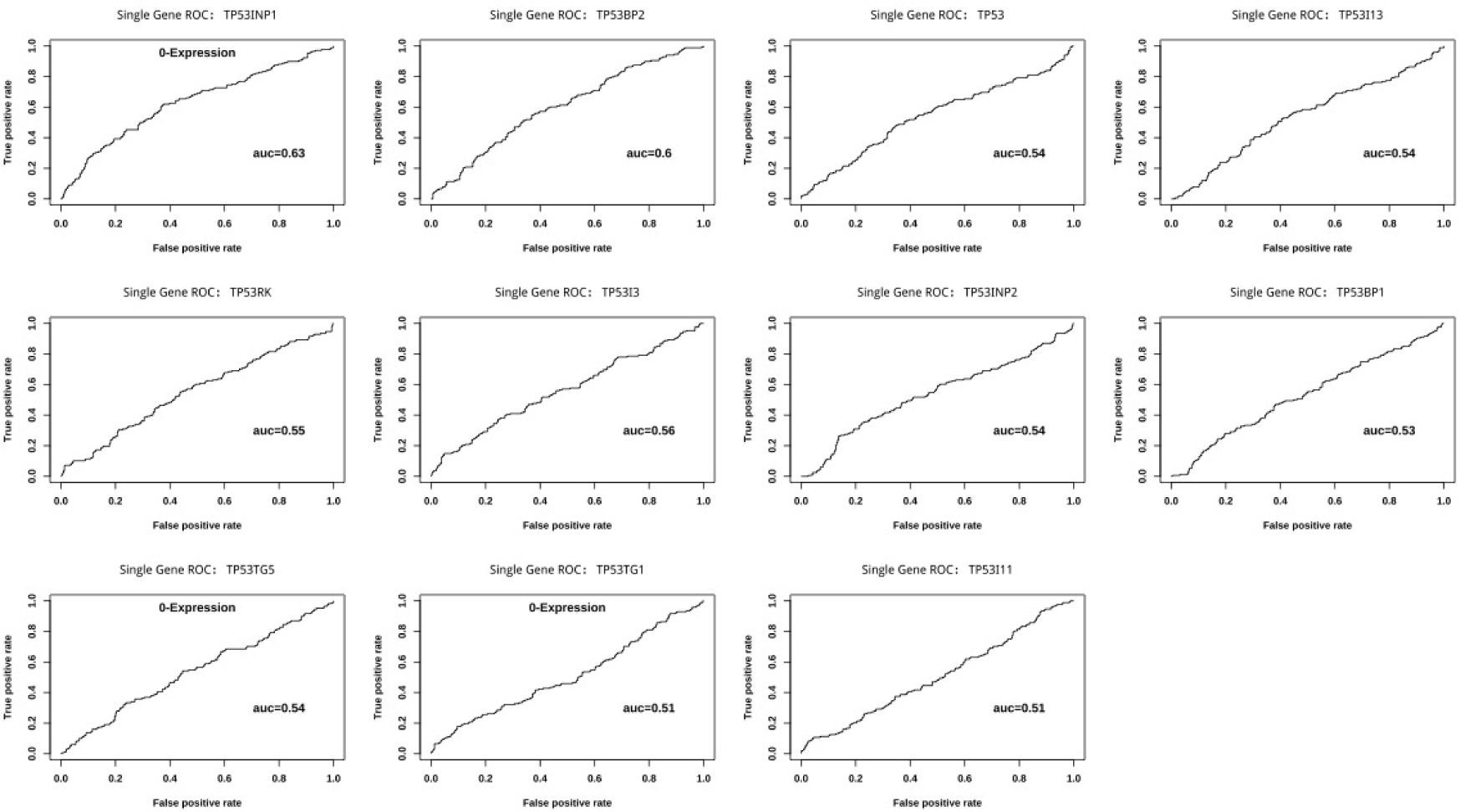
Univariate ROCs of 11 genes in TP53 cluster.

**Table 12:**
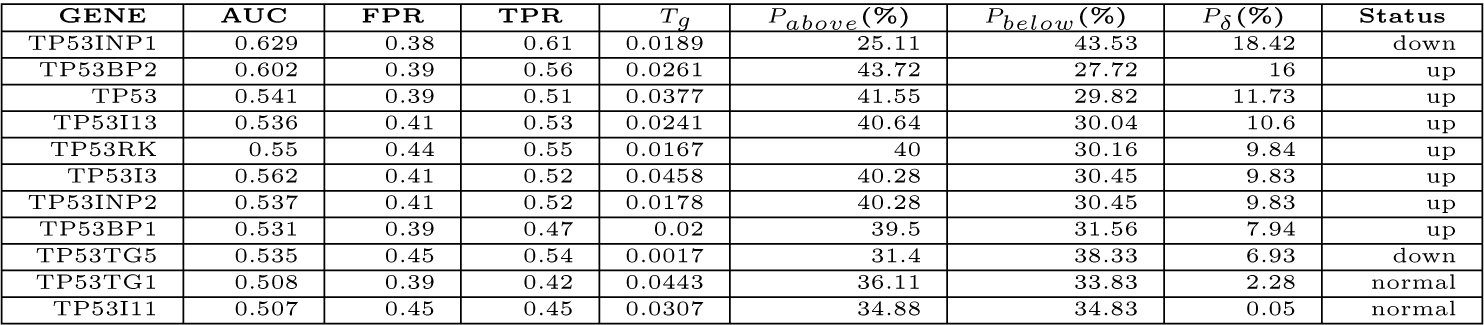
AUCs and recurrence risks of TP53 genes ordered by *P_δ_*.

#### 3.1.10 PDCD1(PD1) Cluster

As shown in Table 1, only 15 genes were pre-selected for PDCD1 cluster. The ROCs are presented in Figure 10 and the corresponding AUCs, FPRs, TPRs, threshold *T_g_*, population risks are listed in Table 13. There are 8 members with *P_δ_ ≥* 5%, accounting for 53%. PTPN11, which is also a member in ROS1 cluster, also appears at the top, PDCD1 suppresses T-cell activation through the recruitment of PTPN11[34]. The second LAG3 and the third PDCD1LG2 have *P_δ_*around 11%. Lymphocyte activation gene 3, LAG3, is a T cell activation inhibitory coreceptors similar to PDCD1 and CTLA4 and emerged as the third important immunotherapy target[35]. LAG3 and PDCD1 synergistically regulate T cell function[66], they collaborate to limit CD8+ T cell signaling and weaken anti-tumor immunity and dual blockade of them is a promising immunotherapy strategy[22]. Moreover, a over-expressed ligand of LAG3, Fibrinogen-like protein 1, FGL1, is also a cluster member and has *P_δ_* = 5.87%. PDCD1LG2 (i.e. PD-L2) is one of two PDCD1 ligands and has been emerged as another immunotherapy target similar to PD-L1[65]. Next tier consists of 3 down-expressed genes with medium prediction power, HLA-DRB1, ZAP70 and PRKCQ, with *P_δ_*range in between 6% and 8.42%. HLA-DRB1 is a HLA Class II Antigen. ZAP70, called Zeta Chain Of T Cell Receptor Associated Protein Kinase, regulates motility, adhesion and cytokine expression of mature T cell. PDCD1 modulation of T cell involves inhibition of TCR-mediated phosphorylation of ZAP70 and association with CD3Z, and downstream inhibition of PKCQ which is required for T cell IL-2 production[53]. Lastly, CD80 with *P_δ_* = 5.85 is also over-expressed. CD80 is a ligand of CTLA4, just like PD-L1 as a ligand of PD1, CD80 and PD-L1 interaction suggests significant crosstalk between PD1 pathways and CTLA4 pathways[51, 75]. However, PD1 itself, CD274/PD-L1, and other important PD1 related gene HLA-DQB1, CD3D/E, CD247(CD3Z), and CD4 have *P_δ_ <* 5, with expression levels not strongly related to lung cancer recurrence by this training data set.

**Figure 10:**
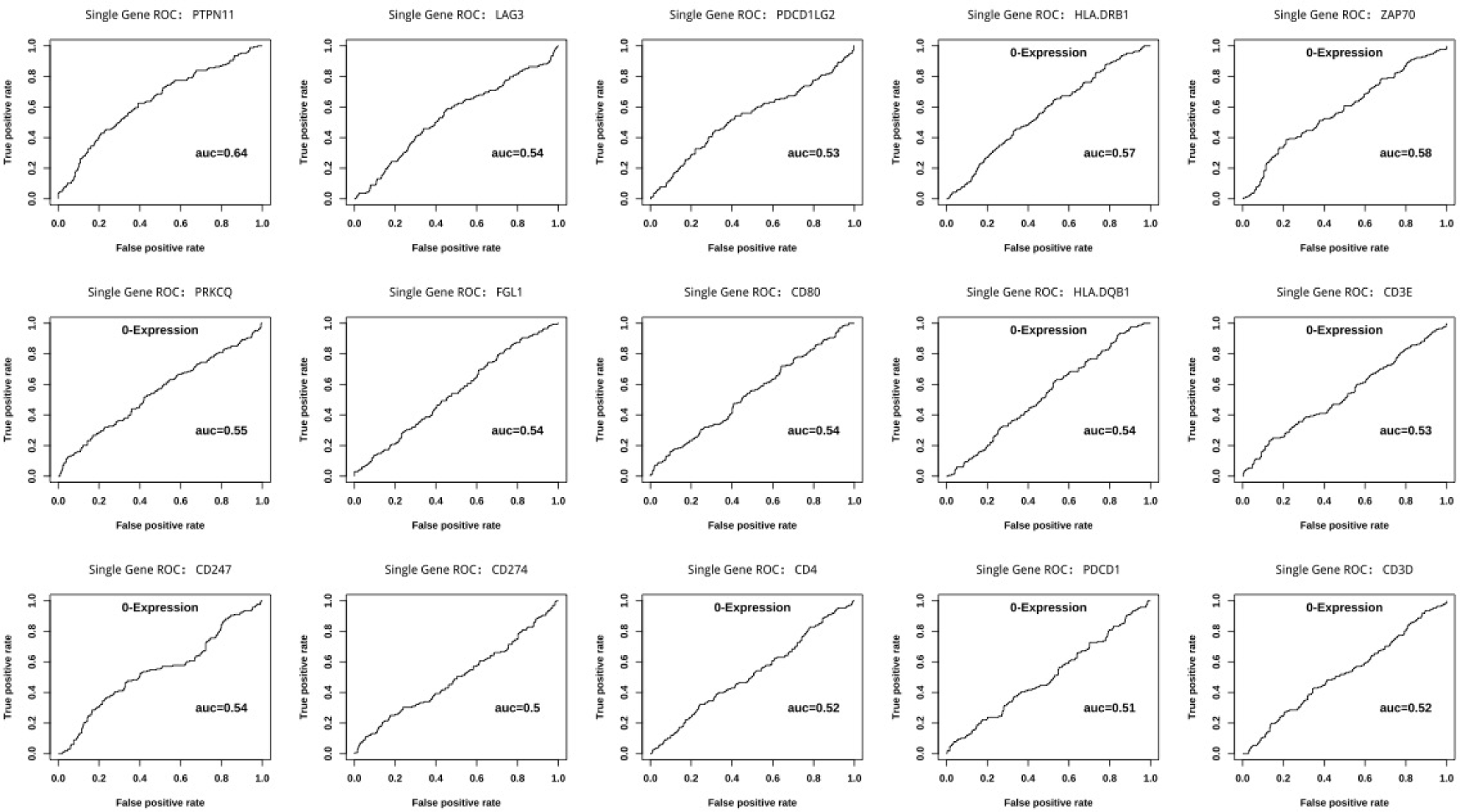
Univariate ROCs of 15 genes in PDCD1 cluster.

**Table 13:**
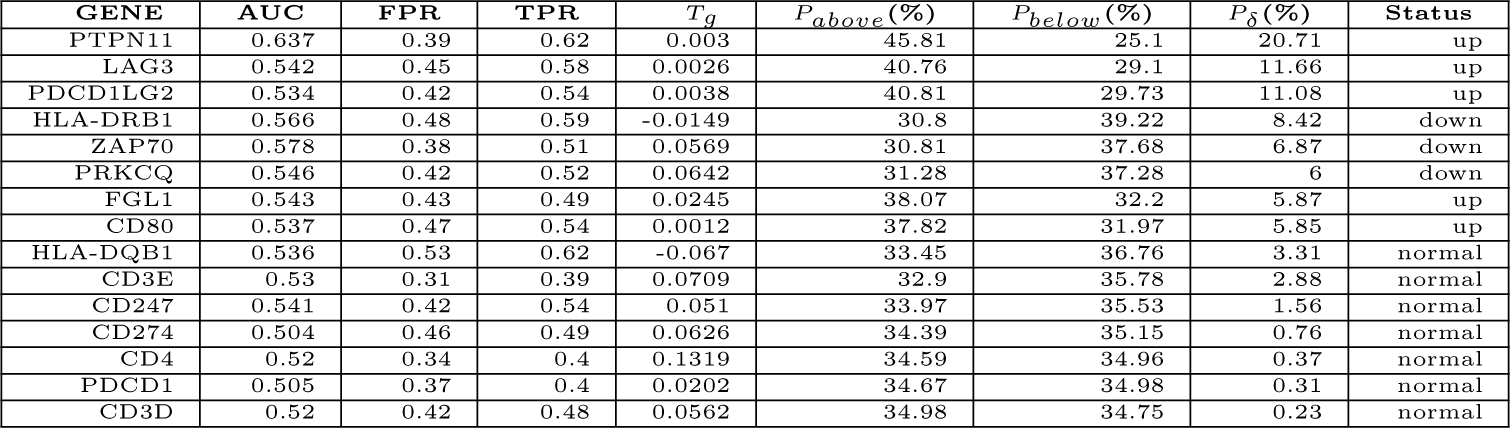
AUCs and recurrence risks of PDCD1 genes ordered by the prediction power

| GENE | AUC | FPR | TPR | $T_g$ | $P_{above}(\%)$ | $P_{below}(\%)$ | $P_\delta(\%)$ | Status |
| --- | --- | --- | --- | --- | --- | --- | --- | --- |
| PTPN11 | 0.637 | 0.39 | 0.62 | 0.003 | 45.81 | 25.1 | 20.71 | up |
| LAG3 | 0.542 | 0.45 | 0.58 | 0.0026 | 40.76 | 29.1 | 11.66 | up |
| PDCD1LG2 | 0.534 | 0.42 | 0.54 | 0.0038 | 40.81 | 29.73 | 11.08 | up |
| HLA-DRB1 | 0.566 | 0.48 | 0.59 | -0.0149 | 30.8 | 39.22 | 8.42 | down |
| ZAP70 | 0.578 | 0.38 | 0.51 | 0.0569 | 30.81 | 37.68 | 6.87 | down |
| PRKCQ | 0.546 | 0.42 | 0.52 | 0.0642 | 31.28 | 37.28 | 6 | down |
| FGL1 | 0.543 | 0.43 | 0.49 | 0.0245 | 38.07 | 32.2 | 5.87 | up |
| CD80 | 0.537 | 0.47 | 0.54 | 0.0012 | 37.82 | 31.97 | 5.85 | up |
| HLA-DQB1 | 0.536 | 0.53 | 0.62 | -0.067 | 33.45 | 36.76 | 3.31 | normal |
| CD3E | 0.53 | 0.31 | 0.39 | 0.0709 | 32.9 | 35.78 | 2.88 | normal |
| CD247 | 0.541 | 0.42 | 0.54 | 0.051 | 33.97 | 35.53 | 1.56 | normal |
| CD274 | 0.504 | 0.46 | 0.49 | 0.0626 | 34.39 | 35.15 | 0.76 | normal |
| CD4 | 0.52 | 0.34 | 0.4 | 0.1319 | 34.59 | 34.96 | 0.37 | normal |
| PDCD1 | 0.505 | 0.37 | 0.4 | 0.0202 | 34.67 | 34.98 | 0.31 | normal |
| CD3D | 0.52 | 0.42 | 0.48 | 0.0562 | 34.98 | 34.75 | 0.23 | normal |

#### 3.1.11 CTLA4 Cluster

CTLA4 cluster contains 17 members. The ROCs are presented in Figure 11 and the corresponding AUCs, FPRs, TPRs, threshold *T_g_*, population risks are listed in Table 14. There are 10 members with *P_δ_≥* 5%, accounting for 59%. Similar to PD1, PTPN11 is still the one with maximal *P_δ_*. CD276 and CD86 are the next highest two over-expressed genes with *P_δ_* very close to PTPN11. CTLA4 itself, CD80 and GRB2 are over-expressed with modest *P_δ_*. CD276, also known as B7-H3, CD80 and CD86 belong to the same B7 family as PD-L1. CTLA4 is a homologue of CD28 and they are coreceptors, GRB2, called growth factor receptor-bound protein 2, is an important adaptor participating CD28 and CTLA-4 signaling mechanisms[49]. CD80/86 binds to CD28 while CTLA4 reduce their interaction time. Synergistically with CTLA4, CD276 inhibits T cell activation by inhibiting IL-2 secretion and evidence suggested that IL20RA is a receptor of CD276[31].

**Figure 11:**
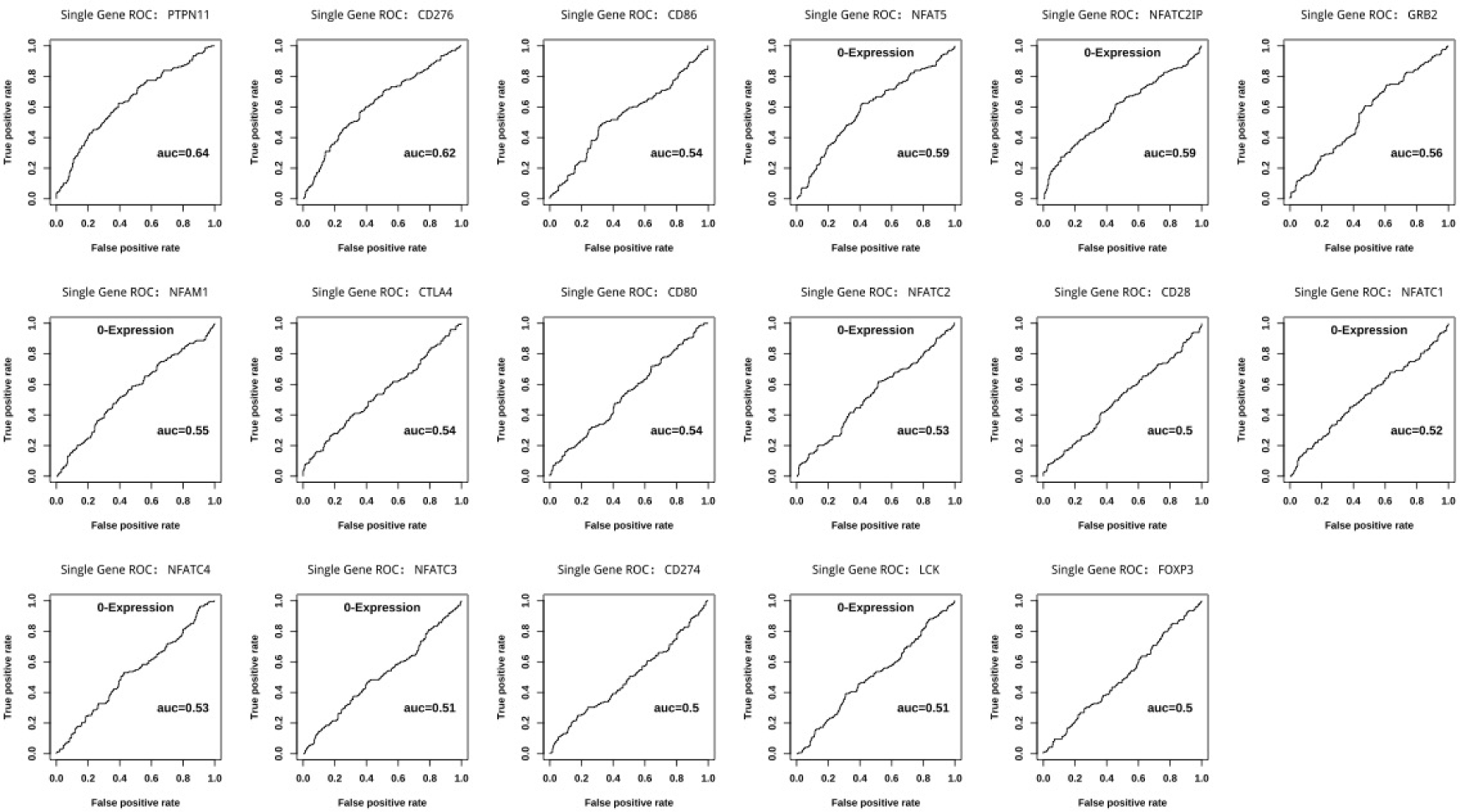
Univariate ROCs of 17 genes in CTLA4 cluster.

**Table 14:**
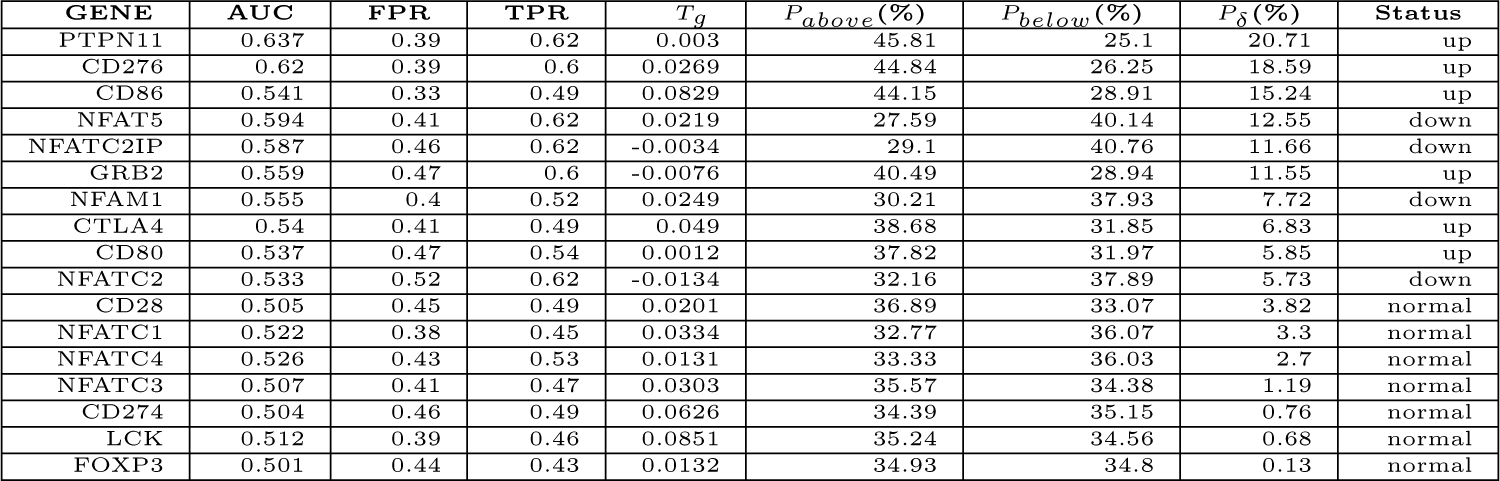
AUCs and recurrence risks of CTLA4 genes ordered by *P_δ_*.

#### 3.1.12 Cluster Member Voting Models

Now that a sample is assigned a percentage of *abnormal* members for each given cluster, another ROC is plotted using the percentage as a recurrence predictor. The ROCs are presented in Figure 12. Table 15 lists the corresponding AUCs, FPRs, TPRs, threshold *T_p_*, *P_above_* representing the recurrence risk of the patient group with abnormal cluster members *≥ T_p_*%, and *P_below_* representing that of the opposite group with *< T_p_*%. In summary, for each cluster, the recurrence risk of the abnormal group (of all pathological stages) ranges from 74% (PDCD1) to 220% (ALK) higher, comparing to the opposite normal group, which is calculated via 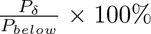. Next the recurrence risks are investigated in more details.

#### 3.1.13 Clusters Defined Using Combinatory GCEI (cGEI)

Given an ordered list of gene clusters represented by ALK, BRAF, EGFR, MET, NTRK, RAS, RET, ROS1 and TP53 in the fixed order, a patient is labeled as a 9-digit binary string *i*_1_*i*_2_ *· · · i*_9_, each digit *i_k_*(*k* = 1,> 2, *· · ·,* 9) stands for the corresponding gene cluster expression status where 0 is for *normal* while 1 for *abnormal*. This is called driver gene cluster expression signature, for example, 000000000 represents that all 9 clusters are *normal*, 100000000 represents that only cluster ALK is *abnormal* while 111111111 represents that all 9 clusters are *abnormal*, etc. The 9-driver gene cluster expression signature classifies lung cancers into 512 (= 2^9^) expression types. Similarly, two immunotherapy target genes: PDCD1, CTLA4 give rise to a two-bit signature string. Moreover, by counting the number of 1 in the 9-digit signature string, which is the number of abnormally expressed clusters in 9 driver gene clusters, called a combinatory *GCEI* and denoted as *cGCEI*. Patients were then grouped into 10 groups with *cGCEI* = 0,> 1,> 2,> 3, *· · ·,* 9 respectively. For example, *cGCEI* = 0 is the patients with signature 000000000, *cGCEI* = 1 is the patients with any signature with only one 1 and eight 0, such as 100000000,> 010000000, *· · ·,* 000000001), and *cGCEI* = 9 is the patients with signature 111111111, etc. Similarly a two-bit binary string by combining GCEI of PDCD1 and CTLA4 is defined and has 3 status: 0, 1, or 2, representing none of, or one of, or both of PDCD1 and CTLA4 clusters are *abnormal*. Furthermore, another combinatory *GCEI* is defined by thresholding *cGCEI* values, a meaningful threshold value of 5 is used to collapse 10 groups with *cGCEI* = 0,> 1,> 2,> 3, *· · ·,* 9 to only two groups, denoted as *DGCntGT* 5, of which the value 1 stands for count of abnormal driver gene cluster is *>* 5 and 0 for *≤* 5. Therefore, *DGCntGT* 5 = 1 means that there are at least 6 abnormal clusters in 9 driver gene clusters, and *DGCntGT* 5 = 0 means that there are at most 5 abnormal driver gene clusters. All these labeling schemes for lung cancers have dramatic indication for recurrence risks, of which the *abnormal* group is in general 130% *−*300% of the *normal* counterpart as shown below.

**Figure 12:**
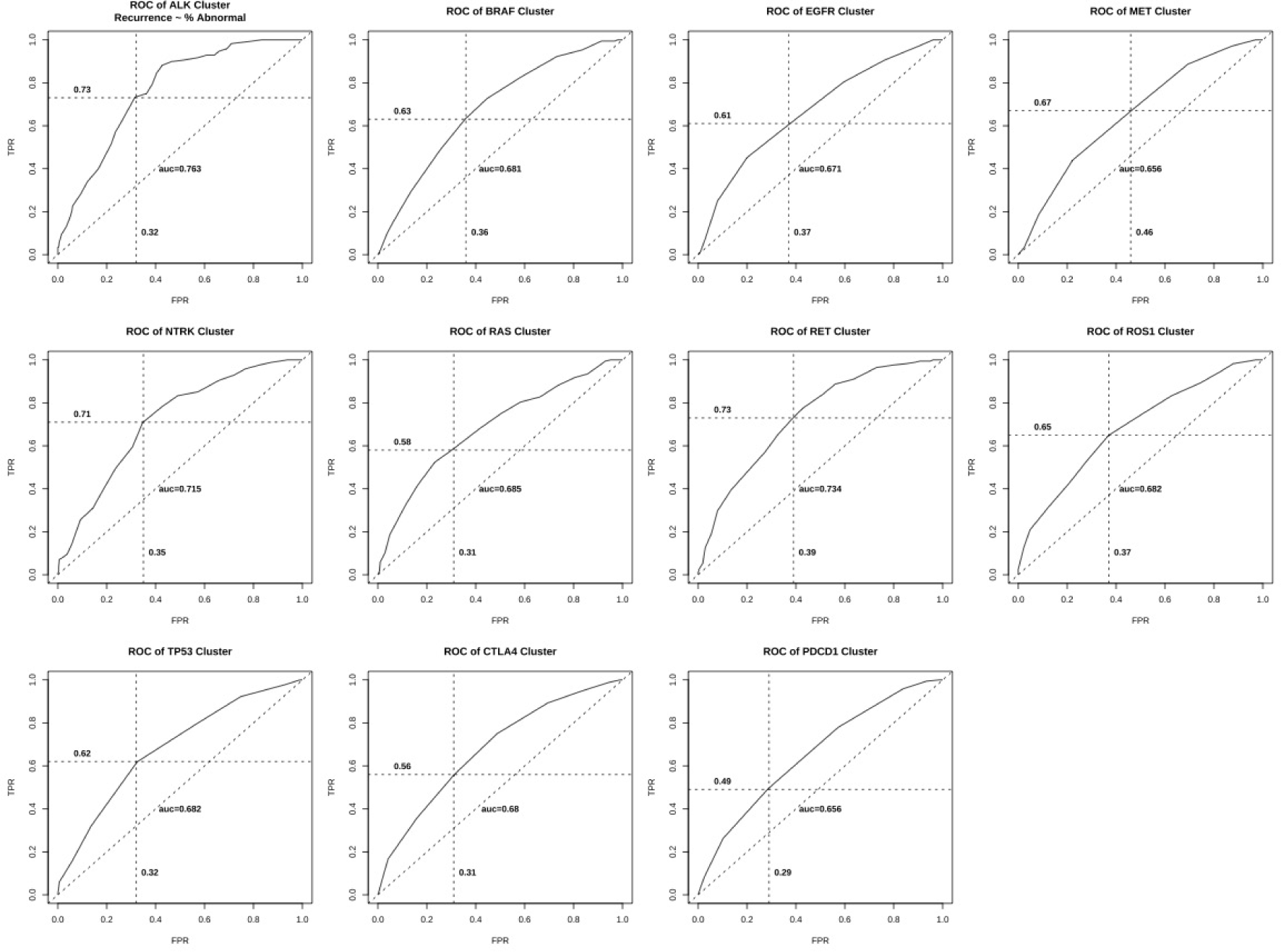
Univariate ROCs of 11 clusters. The percentage of *abnormal* members in each cluster was used as a predictor to recurrence.

**Table 15:** AUC, TPR, FPR, Threshold *T_p_*and Recurrence Risks for 11 Clusters. *P_above_*is the recurrence risk of the patients with the corresponding abnormal cluster members *≥ T_p_*%, and *P_below_* is the opposite group with *< T_p_*%.

| SEED | AUC | $T_p(\%)$ | $P_{above}(\%)$ | $P_{below}(\%)$ | $P_\delta(\%)$ | FPR | TPR | ACCURACY | PPV |
| --- | --- | --- | --- | --- | --- | --- | --- | --- | --- |
| ALK | 0.763 | 55.56 | 55.41 | 17.31 | 38.1 | 0.32 | 0.73 | 0.7 | 0.55 |
| BRAF | 0.681 | 57.89 | 48.62 | 23.48 | 25.14 | 0.36 | 0.63 | 0.64 | 0.49 |
| EGFR | 0.671 | 58.33 | 46.58 | 25.1 | 21.48 | 0.37 | 0.61 | 0.62 | 0.47 |
| MET | 0.656 | 57.14 | 43.75 | 24.78 | 18.97 | 0.46 | 0.67 | 0.59 | 0.44 |
| NTRK | 0.715 | 51.35 | 52.19 | 19.29 | 32.9 | 0.35 | 0.71 | 0.67 | 0.52 |
| RAS | 0.685 | 60 | 50.52 | 24.31 | 26.21 | 0.31 | 0.58 | 0.66 | 0.51 |
| RET | 0.734 | 55.32 | 50.21 | 19.25 | 30.96 | 0.39 | 0.73 | 0.65 | 0.5 |
| ROS1 | 0.682 | 52.38 | 48.44 | 22.96 | 25.48 | 0.37 | 0.65 | 0.64 | 0.48 |
| TP53 | 0.682 | 55.56 | 50.49 | 23.19 | 27.3 | 0.32 | 0.62 | 0.66 | 0.5 |
| CTLA4 | 0.68 | 60 | 48.96 | 25.52 | 23.44 | 0.31 | 0.56 | 0.64 | 0.49 |
| PDCD1 | 0.656 | 62.5 | 47.98 | 27.51 | 20.47 | 0.29 | 0.49 | 0.64 | 0.48 |

### 3.2 Recurrence Risks

In the above lung cancers were labeled as *normal* (*GCEI* = 0) or *abnormal* (*GCEI* = 1) with respect to a given cluster or a combination of atomic GCEIs. Next the recurrence risks were assessed for the subpopulations defined by individual GCEI status and combinations of GCEIs. For a given atomic or a combinatory GCEI, the recurrence risk, defined as the percentage of the recurred patients, was calculated with respect to the GCEI status for patients of different pathological stages, namely of stage I, of stage II-V, and of all stages respectively. Table 16 lists the recurrence risks for subpopulations labeled by the atomic GCEI indicators and *DGCntGT* 5 indicator. It shows that ALK cluster gives the largest risk ratio of lung cancer group with *GCEI* = 1 over *GCEI* = 0 for 3 stage groups, with 320%,> 332%,> 188% for all stages, Stage I, Stage II-IV respectively. As an example of the ratio calculation, take the values in Table 16 corresponding to ALK for all stages, 320% was derived by 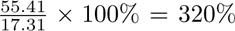, other ratios were calculated similarly. As for the minimal ratio, PDCD1 gives 174% for all stages, MET gives 169% for Stage I, and EGFR gives 109% for Stage II-IV. On average, the risk ratio of group with *GCEI* = 1 over *GCEI* = 0 is 222%,> 247%,> 134% for all stages, Stage I, Stage II-IV respectively. This demonstrates the power of recurrence risk stratification with gene cluster expression voting strategy.

Furthermore, the recurrence risks were also calculated based on binary string signatures of the atomic GCEIs. As described in the above, cGCEIs corresponding to the ordered 9-gene list (ALK, BRAF, EGFR, MET, NTRK, RAS, RET, ROS1, TP53) seperate lung cancers into 10 groups by counting number of *abnormal* clusters, or the number of 1 in the signature string. Table 17 listed the recurrence percentages for 10 groups defined by 9-gene signatures. It shows that the recurrence risk increases along with cGCEI values, namely the number of *abnormal* clusters. For *cGCEI* = 0, the recurrence risk is merely 7.02%, when there is one and only one abnormal cluster (*cGCEI* = 0), the risk more than doubled to 15.28%, and then increases to 20.41% for *cGCEI* = 3. Interestingly, it then comes a hiccup where the risk goes down to 17.50% for *cGCEI* = 4, but this might be due to the data size. After *cGCEI ≥* 6, the risk goes beyond 56.36% to an astonish 72.73% for the group of patients with *cGCEI* = 9 where all 9 diver clusters show abnormal expressions based on member voting and it has only one signature 111111111. This explains the rational that we defined a new GCEI based on *DGCntGT* 5 in the proceeding sub-section.

**Table 16:** Recurrence Percentages of Lung Cancers in Different Stage Groups Flagged by GCEI. Group risk of *GCEI* = 1 is typically 130% to 300% of the corresponding group of *GCEI* = 0.

| Subpopulation | All (Stage I-IV) |  | Stage I |  | Stage II-IV |  |
| --- | --- | --- | --- | --- | --- | --- |
|  | GCEI=0(%) | GCEI=1(%) | GCEI=0(%) | GCEI=1(%) | GCEI=0(%) | GCEI=1(%) |
| ALK | 17.31 | 55.41 | 12.56 | 41.75 | 35.85 | 67.23 |
| BRAF | 23.48 | 48.62 | 15.38 | 36.27 | 53.57 | 59.48 |
| EGFR | 25.1 | 46.58 | 16.67 | 33.02 | 54.24 | 59.29 |
| MET | 24.78 | 43.75 | 17.14 | 28.89 | 50.98 | 60.33 |
| NTRK | 19.29 | 52.19 | 12.87 | 39.81 | 44.23 | 63.33 |
| RAS | 24.31 | 50.52 | 16.36 | 35.42 | 47.3 | 65.31 |
| RET | 19.25 | 50.21 | 14.14 | 35.29 | 39.58 | 64.52 |
| ROS1 | 22.96 | 48.44 | 16.92 | 32.11 | 44.64 | 63.79 |
| TP53 | 23.19 | 50.49 | 14.56 | 37.5 | 48.57 | 63.73 |
| CTLA4 | 25.52 | 48.96 | 13.79 | 38.32 | 52.87 | 62.35 |
| PDCD1 | 27.51 | 47.98 | 16.13 | 36.56 | 54.35 | 61.25 |
| DGCntGT5 | 18.84 | 59.47 | 13.3 | 49.35 | 40.68 | 66.37 |
| Average | 22.63 | 50.22 | 14.98 | 37.02 | 47.24 | 63.08 |

**Table 17:** Recurrence Risks of cGCEI based on 9-digit Signatures (Only Evaluated for all Stages).

| cGCEI | Ex Signature | NoneRecurred | Recurred | Total | Recurrence(%) |
| --- | --- | --- | --- | --- | --- |
| 0 | 000000000 | 53 | 4 | 57 | 7.02 |
| 1 | 000000001 | 61 | 11 | 72 | 15.28 |
| 2 | 000001100 | 39 | 10 | 49 | 20.41 |
| 3 | 000100101 | 33 | 7 | 40 | 17.5 |
| 4 | 101100010 | 30 | 12 | 42 | 28.57 |
| 5 | 001011011 | 21 | 11 | 32 | 34.38 |
| 6 | 111110001 | 24 | 31 | 55 | 56.36 |
| 7 | 111110101 | 24 | 26 | 50 | 52 |
| 8 | 111110111 | 20 | 32 | 52 | 61.54 |
| 9 | 111111111 | 9 | 24 | 33 | 72.73 |

## 4 Discussion

Although DNA-based genetic tests have been routinely used for targeted therapy and immunotherapy, the proportion of patients whose tumors can be targeted therapeutically is limited and is usually less than 30%. A retrospective study of 2257 metastatic NSCLC patients showed that more than half of tested patients did not have results prior to first-line treatment and fewer than 20% of tested patients had results for all 4 driver mutations (ALK, EGFR, ROS1, BRAF) and PD-L1 prior to first-line treatment. Moreover, although the turnaround time improved from year 2017 to 2019, not all patients who tested positive for driver mutations received targeted therapy in the first-line setting[37]. Hence the percent of patients who received targeted therapy was less than 30%. We propose that for a given driver gene cluster, the targeted therapy with respect to the gene may be beneficial to the patient group of *GCEI* = 1. In addition, immunotherapy may be beneficial to the patient group of *GCEI* = 1 with respect to PDCD1 or CTLA4 clusters. The WINTHER trial (NCT01856296) [47] was the first clinical trial to navigate lung, colon, head and neck, and other cancer patients with previous treatments to therapy on the basis of fresh biopsy-derived DNA sequencing or RNA expression (tumor versus normal). It shows that transcriptome profiling is as useful as DNA tests for improving therapy recommendations and patient outcome, and hence transcriptome analysis can expand personalized treatment.

## Data Availability

All data produced in the present study are available upon reasonable request to the authors.

https://www.ncbi.nlm.nih.gov/geo/query/acc.cgi?acc=GSE30219

https://www.ncbi.nlm.nih.gov/geo/query/acc.cgi?acc=GSE31210

## Abbreviations

GCEI: Gene Cluster Expression Index
GEO: Gene Expression Omnibus
ROC: Receiver Operating Curve
AUC: Area Under the Curve
FPR: False Positive Rate
TPR: True Positive Rate
PPV: Positive Prediction Value

